# Cerebral Cortical Reorganization After Intracerebral Hemorrhage in Children

**DOI:** 10.64898/2025.12.19.25341480

**Authors:** Hannah L. Choi, Shivani Mahuvakar, Jonas Schollenberger, Rebecca Shuere, Amir Sadikov, Lanya T. Cai, Christine Mrakotsky, Kathryn Nesbit, Laura G. Hess, Helen Kim, David A. Saloner, Christine K. Fox, Jarod L. Roland, Heather J. Fullerton, Pratik Mukherjee, UCSF AHA Bugher Hemorrhagic Stroke Research Group

## Abstract

**BACKGROUND AND PURPOSE:** Structural changes following pediatric intracerebral hemorrhage (ICH) caused by ruptured brain vascular malformations remain poorly understood. We conducted a longitudinal study to examine morphometric changes in brain volume and cortical thickness across ipsilesional and contralesional hemispheres following unilateral ICH.

**METHODS:** Brain magnetic resonance imaging (MRI) was acquired in 20 patients aged 6–18 years (*Median:* 13, *IQR:* 9.3–16) at presentation of ICH prior to treatment of the hemorrhage etiology (baseline/Session 1) and repeated at 6 months (Session 2) and 12 months (Session 3) post-ICH. T1- and T2-weighted Fluid-Attenuated Inversion Recovery (T2-FLAIR) MR images were segmented into cortical and subcortical regions, with gray matter parcellated based on the Desikan-Killiany-Tourville (DKT) atlas. We used deep learning-based lesion extraction to enable more precise measurements of brain volume and cortical thickness. Morphometric changes were quantified as percentage change from baseline with signed values denoting increase or decrease. We analyzed longitudinal change in volume and cortical thickness of contralesional and ipsilesional hemispheres and lobes. Structural covariance analyses focused on correlations of thickness between homologous and non-homologous cortical regions. Significant correlations were classified based on the sensorimotor–association axis, Mesulam laminar differentiation, DKT regions, von Economo-Koskinas cytoarchitecture, and Yeo 7-network functional MRI (fMRI) atlases.

**RESULTS:** Pediatric post-ICH recovery was characterized by dynamic and spatially divergent morphometric changes. At 6 months, contralesional cortical regions exhibited increases in volume and thickness; however, by 12 months, ipsilesional cortical, subcortical, and white matter structures showed progressive reductions. Morphological reorganization was prominent in the contralesional hemisphere; association, heteromodal, and frontal cortices; and within default mode and frontoparietal functional networks.

**CONCLUSIONS:** Our findings highlight the vulnerability and plasticity of higher-order cortical regions and implicate contralesional cortical networks as substrates of resilience in recovery.

## INTRODUCTION

Intracerebral hemorrhage (ICH) is a form of hemorrhagic stroke characterized by the abrupt onset of bleeding within the brain parenchyma. Subarachnoid hemorrhage, the other main type of hemorrhagic stroke, involves bleeding into the subarachnoid space; however, our study concentrates on pediatric ICH. In pediatric populations, hemorrhagic strokes account for 39% to 54% of all strokes (the remainder being ischemic strokes)^1–3^. In contrast, among adults, hemorrhagic strokes constitute only 7.5% to 9% of all strokes^1,4^. Children present differently from adults in various stroke types, yet research on pediatric stroke largely mirrors adult research trends prioritizing ischemic strokes^1,5^. This is particularly concerning given that hemorrhagic stroke presents a notably higher risk of mortality than ischemic stroke in both children^6–8^ and adults^4,9^. For children, mortality rates from hemorrhagic stroke are approximately double those of ischemic stroke^6–8^. Furthermore, many studies report that around a quarter to a half of children who survive a hemorrhagic stroke experience long-term functional impairments such as motor and cognitive deficits^10–13^ that may eventually lead to compromised independent living later in life^10,14^. Altogether, further research is needed to better understand pediatric ICH as a foundation of optimizing patient outcomes. Our longitudinal study examining the structural changes following pediatric ICH aims to contribute to this effort by further elucidating brain recovery post-ICH.

The response of the pediatric brain to ICH remains under-investigated, particularly the progression of cerebral changes occurring during stroke recovery. We specifically examine unilateral ICH resulting from ruptured cerebral vascular malformations due to arteriovenous malformations (AVMs), cavernous malformations (CMs), and aneurysms. AVMs are the most common cause of pediatric ICH, occurring at a rate 10 times higher than aneurysms^5,12,15^. Additionally, the unilateral presentation of ICH in our cohort allows for an examination of the inter-hemispheric, or transcallosal, mechanisms of neurological recovery, which may extend to a broader spectrum of unilateral cerebral pathologies.

Our study aims to address important gaps in understanding post-ICH recovery by adopting a longitudinal approach, acquiring imaging at 6-month intervals following the onset of pediatric ICH and treatment of its etiology, affording a glimpse into ongoing recovery or degeneration over successive months. The longitudinal design is especially important in children and adolescents, a population with wide age-related variation in brain morphological metrics. We utilize modern neuroimaging techniques that facilitate more precise lesion segmentation and whole-brain structural analysis, specifically volumetric and cortical thickness measurements, through state-of-the-art segmentation and parcellation tools^16^. Our experimental framework, alongside a distinctive dataset spanning 12 months for each individual case of pediatric ICH, as well as contemporary analysis software, uniquely positions us to investigate recovery trajectories and consequently identify pivotal time windows which may shed light on the rate and pattern of recovery from pediatric ICH.

We hypothesize that damage is not confined to the affected hemisphere but may have transcallosal effects, or contralesional involvement, due to the interconnectivity of brain regions^17–22^. Thus, it is anticipated that damage occurring in one hemisphere (ipsilesional) will have a counterpart in the opposite hemisphere (contralesional), and that positive neuroplastic mechanisms observed in one side of the brain are expected to be mirrored on the other. Additionally, while a majority of cortical alterations are anticipated in the ipsilesional hemisphere and contralesional homologous areas to a lesser degree^23–27^, significant variations in structural metrics are also expected in remote regions^28–30^, reflecting network-mediated distant effects of injury and recovery through diaschisis and recruitment, respectively. However, changes in morphometry of cerebral cortex (i.e., volume or thickness) over time may present ambiguities, indicating either neuronal effects (i.e., hypertrophy/atrophy, synaptogenesis/synaptic pruning), or, conversely, conditions such as vasogenic edema, resolution of edema, and/or neuroinflammation. Lastly, more structural changes are expected to occur in the early stages of post-ICH recovery than in the later stages^31^. Through this research, we seek to expand our understanding of the neurobiological processes guiding recovery in pediatric ICH, and perhaps more generally in unilateral cerebral pathologies, providing insights that could contribute to future therapeutic interventions such as image-guided neuromodulation.

## MATERIALS AND METHODS

### Participants

Our single-center prospective cohort study recruited children and adolescents who were diagnosed with unilateral ICH, resulting from ruptured cerebral vascular malformations, including AVMs, CMs, and aneurysms, in accordance with a research protocol approved by the institutional review board at our medical center. In this study, ICH is defined as a focal collection of blood within the brain parenchyma that is not caused by trauma or by hemorrhagic transformation of an arterial or venous infarct. Consent from parents or guardians, and assent from patients when appropriate, were obtained. Investigators and clinicians ensured that participants met the necessary qualifications for additional research sequences, prioritizing safety concerns and data quality. Finally, pregnancy resulted in ineligibility to participate. Please refer to the supplemental material for a detailed list of the study’s inclusion and exclusion criteria (Table S1).

### Experimental Design and MRI Data Acquisition

Magnetic resonance imaging (MRI) scans were acquired over 3 sessions using a GE 3-Tesla Discovery MR750 scanner. The initial session (Session 1), serving as a baseline, took place shortly before the treatment of the hemorrhage etiology, which included embolization, surgical resection, or gamma knife therapy. Subsequent scans were performed at 6 ± 3 months (Session 2) and 12 ± 3 months (Session 3) after ICH (Fig. 1). 3D T1-weighted (T1w) gradient echo imaging and 3D T2-weighted Fluid-Attenuated Inversion Recovery (T2-FLAIR) imaging were performed, both at 1mm isotropic spatial resolution.

**Figure 1.**
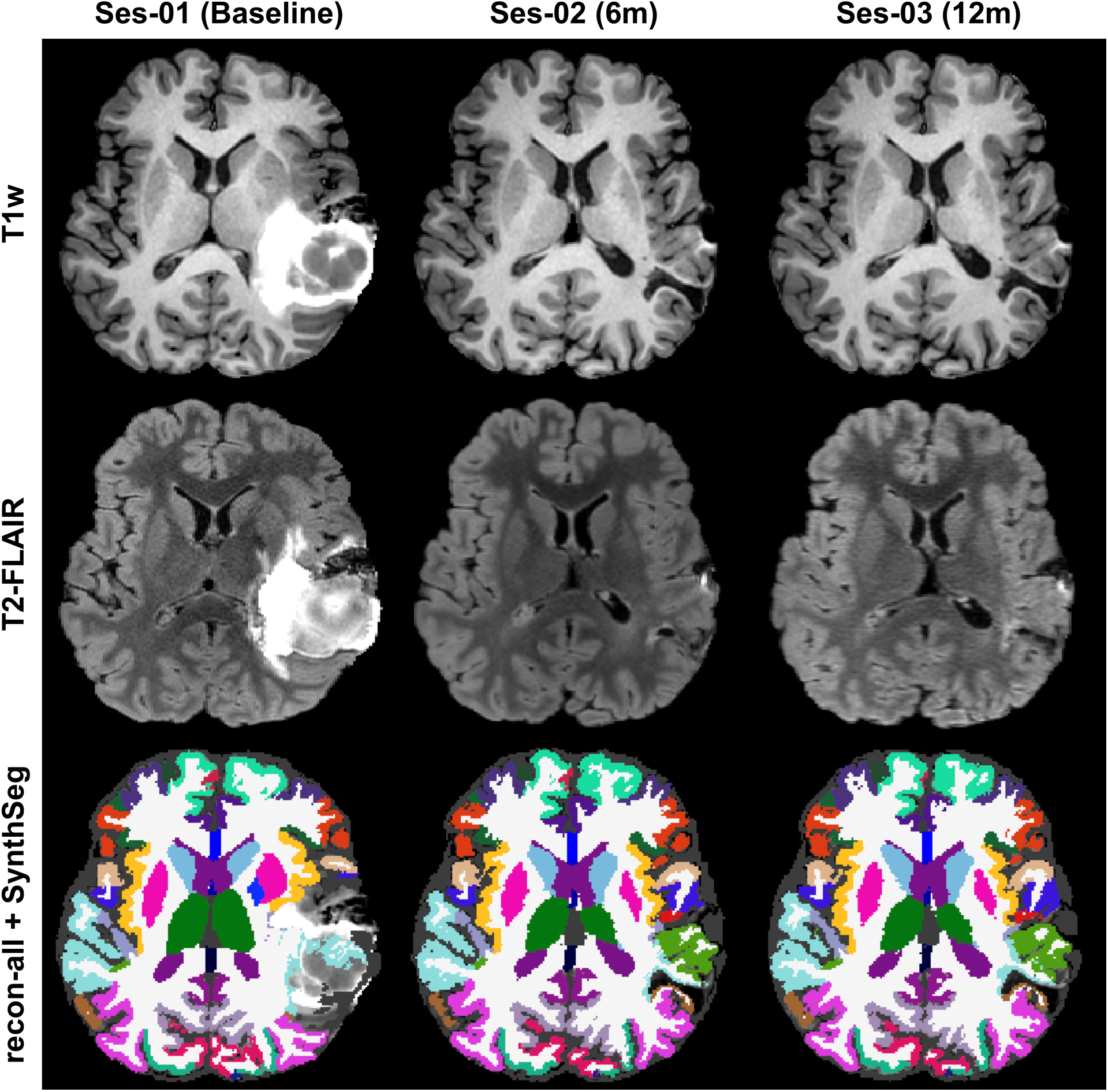
Longitudinal brain imaging at 6-month intervals. (*Top*) T1w, (*Middle*) T2-FLAIR, and (*Bottom*) parcellated MRI with patient-specific DKT atlas regions are shown over 3 timepoints including (*Left*) Session 1 at baseline, (*Center*) Session 2 at 6 months post-ICH, and (*Right*) Session 3 at 12 months post-ICH.

### MRI Data Preprocessing

#### Anatomical Segmentation and Parcellation

All MRI scans were carefully inspected to confirm the absence of artifacts. We employed Freesurfer 7.4.0, utilizing its recon-all pipeline integrated with SynthSeg^32^. T1w scans were always incorporated, and when T2-FLAIR images were available, they were additionally used to enhance the delineation of pial surfaces (Fig. 1). Post-segmentation and parcellation, we conducted a second thorough visual inspection of all the processed images to ensure that the hemorrhage and brain atrophy were appropriately accounted for. We include Dice scores indicating segmentation performance of the select brain regions tested by SynthSeg (Fig. S1). We referred to the Desikan-Killiany-Tourville (DKT) Atlas^33^ to group brain regions into lobes and areas, including the frontal, temporal, parietal, and occipital lobes as well as the cingulum and insula^33^.

#### Neuromaps Parcellation

We utilize the neuromaps framework^34^ to examine cortical changes against five macroscale atlases defined by underlying cortical microarchitecture: the sensorimotor–association (SA) axis^35^, Mesulam’s hierarchy of laminar differentiation^36^, DKT atlas^33^, von Economo-Koskinas cytoarchitectural atlas^37^, and Yeo 7-network functional MRI (fMRI) atlas^38^.

The SA axis, a principal gradient of macroscale cortical organization, was used to situate each region along a continuum from primary and specialized sensorimotor regions to higher-order association areas, which encompass the broader abstract, integrative, and affective domains of cognition. Mesulam’s hierarchy of laminar differentiation, positioned along the SA axis, served to further delineate the cortex into four tiers, ranging from: 1) *idiotypic cortex*, which supports primary sensory and motor functions; to 2) *unimodal cortex*, which mediates higher-order processing within a single modality such as vision, touch, or audition; to 3) *heteromodal cortex*, which integrates across multiple sensory, motor, and cognitive domains; and finally to 4) *paralimbic cortex*, where cognitive and affective signals converge to guide behavior.

The DKT atlas was used to parcellate the cortex into 62 cortical regions (31 left, 31 right) that were further grouped into traditional lobes and limbic brain areas: the frontal lobe, the parietal lobe, the occipital lobe, the cingulate cortex, and the insula.

The von Economo-Koskinas atlas was employed to order regions from thinner to thicker cortices, ranging from: 1) *granular cortex (koniocortex)*, marked by layer IV, specialized for sensory processing; 2) *polar cortex*, located at the poles of lobes and containing granule and stellate cell populations; 3) *parietal cortex*, notable for its prominent layers II and IV, composed of small to medium pyramidal cells and inhibitory interneurons; 4) *frontal cortex*, characterized by larger but less densely packed pyramidal neurons than in parietal cortex; and 5) *agranular cortex*, which features a sparse layer IV but prominent layers III and V with large pyramidal neurons involved in sensorimotor integration and motor planning/execution.

Finally, the Yeo parcellation from resting state fMRI was utilized to delineate seven canonical functional connectivity networks: 1) *visual*, which supports the processing of visual information and visuospatial integration; 2) *somatomotor*, which underlies primary sensory and motor functions including voluntary movement and somatosensory processing; 3) *dorsal attention*, which mediates top-down allocation of attention and goal-directed visuospatial processing; 4) *ventral attention*, which detects behaviorally salient events and reorients attention accordingly; 5) *limbic*, which contributes to affective regulation, valuation, and memory-related processes; 6) *frontoparietal*, which underlies flexible cognitive control, working memory, and adaptive behavior across contexts; and 7) *default mode*, which is active when the mind is at rest such as during self-referential thought or thinking about the past.

All neuromaps used in this study were obtained from publicly available packages: the ENIGMA toolbox^39^, the Yeo 7-network atlas^38^ provided in FreeSurfer, and an in-house package made available at https://github.com/ucsfncl/diffusion_neuromaps^40^. We mapped DKT regions to the Yeo networks by calculating the voxel-wise overlap between each DKT region and each Yeo network. A region was assigned to a given network if the combined voxel overlap of its homologous pair exceeded 300 voxels. For all neuromaps, a region-to-region correlation pair (i.e., connection, edge) was classified into a given atlas category if both constituent regions were members of that class. Accordingly, some edges remained unclassified.

### Statistical Analysis

Our MRI data analysis focused on evaluating changes in brain volume and cortical thickness in gray matter (GM) lobes and regions, comparing between the ipsilesional and contralesional hemispheres. In addition, we assessed hemispheric white matter (WM) volume derived from FreeSurfer’s tissue segmentation. The primary metrics employed were the percent change relative to the baseline session (Session 1), offering a measure of morphometric change from the presentation of ICH, and the percent change relative to each prior session, offering a measure of morphometric change between consecutive 6-month timepoints. Percent change relative to baseline was calculated as [(*Session N* – *Session 1*) / *Session 1*] x 100 and as [(*Session N* – *Session N*–*1*) / *Session N*–*1*] x 100 for percent change between sessions for the morphometric indices (i.e., volume, cortical thickness) of each GM lobe or region, yielding signed values that indicate either an increase (+) or a decrease (–) in size. A positive percentage change indicates an increase in size, potentially indicative of edema or growth in GM (e.g., synaptogenesis). Conversely, a negative percent change indicates a decrease in size, pointing towards potential atrophy or local and/or remote degeneration (i.e., diaschisis).

For statistical analysis, group differences were examined using the Mann-Whitney U Test, with Benjamini-Hochberg (BH) corrections to control for multiple comparisons. Additionally, simple linear regression analyses were conducted along with the calculation of Pearson Correlation Coefficients, to assess inter-hemispheric properties, specifically the relationship between morphometric changes in the ipsilesional and contralesional hemispheres, which represented x and y axes, respectively. To substantiate these correlations, permutation testing (*n* = 5000) was performed, offering a robust, non-parametric means to ascertain whether the observed changes in morphometry demonstrate a genuine correlation between homologous over non-homologous lobes and regions. We additionally computed correlations (i.e., connectivity matrices) across homologous and non-homologous regions to identify the strongest and most significant network edges, assessed globally across the brain.

Normalization of brain volume was performed such that each participant’s brain area, lobe, or region was divided by their corresponding estimated total intracranial volume (eTIV), which is provided by the FreeSurfer recon-all pipeline. These normalized values were then used to compute the percent change relative to the baseline session or between consecutive sessions. In addition, since eTIV is expected to be consistent throughout all sessions and can be utilized as a quality control metric of proper brain segmentation, we conducted further longitudinal examination of the eTIV of participants over sessions and excluded participants who deviated more than 5 percent from baseline from the normalized analyses presented in the supplemental material.

Finally, all analyses and visualizations were performed in Python version 3.13.1^41^ and R version 4.4.2^42^. For visualization, we used the ENIGMA Toolbox^39^, Nilearn^43^, pyCirclize^44^, Seaborn^45^, and Matplotlib^46^ in Python and ggplot2^47^ in R. The formatting of the circular connectograms was adapted from Daianu et al. (2016)^48^. For statistical analyses, we used NumPy^49^, pandas^50^, scikit-learn^51^, SciPy^52^, statsmodels^53^, and pingouin^54^ in Python and tidyr^55^ and dplyr^56^ in R.

## RESULTS

### Patient Demographics, Lesion Characteristics, and Treatment Details

We identified 20 participants, of whom 16 underwent three consecutive structural brain scans and 4 underwent two. The age range of the participants was 6 to 18 years (*Median:* 13, *IQR:* 9.3–16). The sex distribution comprised 11 females (55%) and the remainder were males.

Eighteen participants (90%) experienced ICH in the left cerebral hemisphere, with a marked predominance in the temporal region (*n* = 10, 50%). The primary pathology involved brain AVMs (*n* = 17, 85%), with surgical resection being the main treatment modality (*n* = 15, 75%). Refer to Tables 1 and S2 for comprehensive details on patient demographics, lesion characteristics, and treatment. In terms of ethnicity, 13 participants (65%) identified as Hispanic or Latino, while 7 (35%) did not. For a further breakdown of ethnicity by race, please see Figure S2.

**Table 1.** Summary of Patient Demographics, Lesion Characteristics, and Treatment Type.

| Age |  |
| --- | --- |
| Median (IQR) | 13 (9.3–16) |
| Minimum, Maximum | 6.8, 17.9 |
| Sex |  |
| Female | 11 (55%) |
| Male | 9 (45%) |
| Side of Lesion |  |
| Left | 18 (90%) |
| Right | 2 (10%) |
| Location of Lesion |  |
| Cerebellar | 3 (15%) |
| Frontoparietal | 1 (5%) |
| Insular | 3 (15%) |
| Occipital | 1 (5%) |
| Paracentral | 1 (5%) |
| Parietal | 1 (5%) |
| Perirolandic | 4 (20%) |
| Temporal | 8 (40%) |
| Temporoparietal | 2 (10%) |
| Diagnosis |  |
| Aneurysm | 1 (5%) |
| AVM | 17 (85%) |
| CM | 1 (5%) |
| Unknown VM | 1 (5%) |
| Treatment Type |  |
| Aneurysm Embolization and Clipping | 1 (5%) |
| AVM Embolization | 2 (10%) |
| AVM Resection | 14 (70%) |
| CM Resection | 1 (5%) |
| Gamma Knife | 4 (20%) |
| N/A | 1 (5%) |
| Number of MRI Sessions |  |
| 2 Sessions | 4 (20%) |
| 3 Sessions | 16 (80%) |

*Note.* AVM, arteriovenous malformation; CM, cavernous malformation/cavernoma.

### Estimated Total Intracranial Volume Across Sessions

Following a longitudinal examination of eTIV across sessions per participant, 5 individuals were identified to exceed 5 percent change in eTIV from baseline (Fig. S3). For our supplementary analyses of normalized brain volume, these 5 participants are excluded, making our adjusted Session 2 *n* = 15 and Session 3 *n* = 12, as opposed to our original Session 2 *n* of 20 and Session 3 *n* of 16. Because of a notable drop in *n*, we include the normalized analyses of brain volume in the supplementary material and incorporate them into the Results and Discussion below with caution.

### Whole-Brain Volume and Cortical Thickness Changes After ICH

#### Whole-Brain Volume

Figure 2 shows whole-brain volume divided by hemisphere of cerebral cortex GM, cerebral WM, subcortical GM, and the total GM (cortical and subcortical GM). Notably, the contralesional cerebral GM displays increases in volume at Session 2, 6 months post-ICH [*U* = 100, *p* = 0.004, *d* = –0.48] (Table S3A). Contrastingly, the ipsilesional cerebral GM displays a downward trend, indicative of neurodegeneration, reaching significance at Session 3, in comparison to Session 1. Additionally, the ipsilesional cerebral WM and subcortical GM exhibit a downward trend of neurodegeneration, significant for both Sessions 2 and 3, from 6 months to 12 months post-ICH. The neurodegeneration is sustained from Sessions 2 to 3 when tested against Session 1; however, no significant differences exist between Sessions 2 and 3. On a global level (both hemispheres), the ipsilesional hemisphere’s decreases are more pronounced over the contralesional hemisphere’s increases in volume. Our normalized analysis (Fig. S4, Table S3B) supports these results but highlights the subcortical GM as the primary significant area of ipsilesional neurodegeneration, for both Sessions 2 [*U* = 225, *p* < 0.001, *d* = 1.12] and 3 [*U* = 165, *p* < 0.001, *d* = 1.32], and the cerebral cortex GM as the significant area of contralesional volume increase. A peak in cortical GM increase is observed at Session 2 [*U* = 45, *p* = 0.003, *d* = –0.98]. Overall, there exists a clear dichotomy between the contralesional hemisphere, which exhibits expansion, and the ipsilesional hemisphere, which exhibits shrinkage in brain volume. Also, contralesional expansion, particularly of the cerebral GM, seems to manifest earlier in Session 2 and fade by Session 3, whereas ipsilesional atrophy seems to sustain from Session 2 and become even more prominent by Session 3.

**Figure 2.**
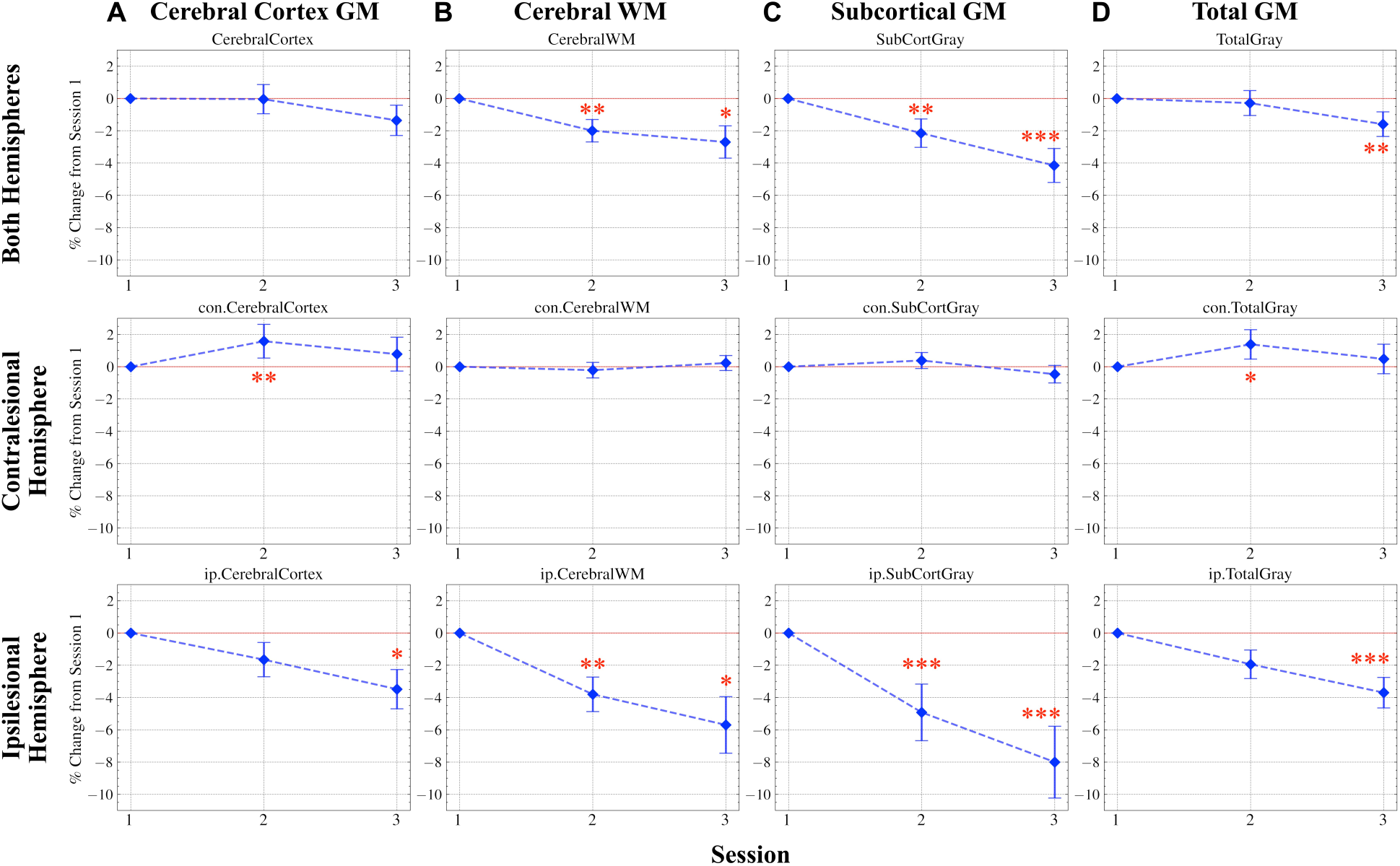
Ipsilesional cerebral GM, WM, and subcortical GM shrinkage and contralesional cerebral GM expansion after pediatric ICH. (**A**) Cerebral Cortex GM, (**B**) Cerebral WM, (**C**) Subcortical GM, and (**D**) Total GM are grouped by (*Top*) Both Hemispheres, (*Middle*) Contralesional Hemisphere, and (*Bottom*) Ipsilesional Hemisphere. * *p* < 0.05, ** *p* < 0.01, *** *p* < 0.001. GM, gray matter; WM, white matter.

#### Whole-Brain Cortical Thickness

Whole-brain cortical thickness divided by hemisphere (Fig. 3, Table S4) reveals a supporting pattern to whole-brain volume (Fig. 2). The mean cortical thickness of the contralesional hemisphere similarly displays an increase for Session 2 [*U* = 100, *p* = 0.004, *d* = –0.33] (Table S4) followed by a return to baseline. The ipsilesional hemisphere displays a decrease particularly in Session 3; however, statistical significance is not reached. Overall, whole-brain cortical thickness reflects the hemispheric divide and temporal dynamics of whole-brain volume changes, with the contralesional cortical thickness peaking at Session 2, in contrast to the ipsilesional cortical thickness plunging at Session 3.

**Figure 3.**
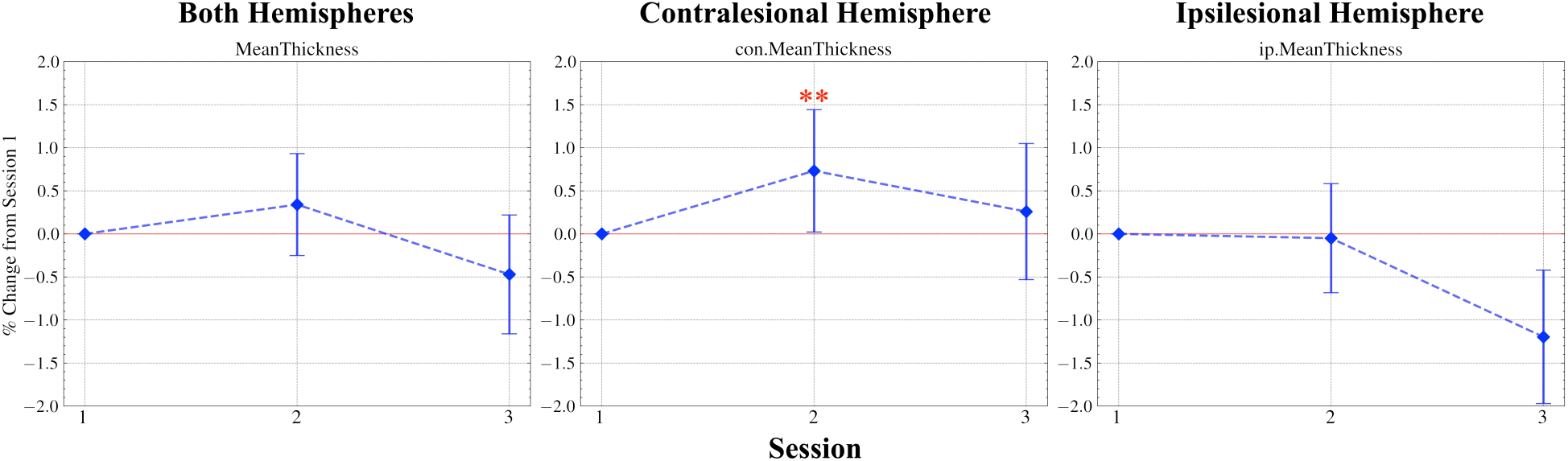
Ipsilesional decrease and contralesional increase of cortical thickness after pediatric ICH. Cortical thickness is grouped by (*Left*) Both Hemispheres, (*Center*) Contralesional Hemisphere, and (*Right*) Ipsilesional Hemisphere. * *p* < 0.05, ** *p* < 0.01, *** *p* < 0.001.

### Lobar and Regional Volume and Cortical Thickness Changes After ICH

#### Lobar Volume

When examining brain volume separated into lobes (Figure 4, Table S5A), it becomes apparent that contralesional cortical GM expansion is widespread and greatest for lobes that encompass or are adjacent to the homologues of lesion sites. We observe some sustainment of volume increase, particularly of the contralesional lateral and medial temporal lobe into Session 3, which are the contralateral homologues to the temporal lobe ICH found in a high proportion of these patients. In contradistinction, the peak increase occurs in Session 2 for the contralesional frontal, parietal, and occipital lobes, followed by a return to baseline in Session 3. The ipsilesional hemisphere displays a contrasting picture again with opposing directionality in volume changes towards atrophy. Specifically, the ipsilesional parietal lobe exhibits the greatest decrease in volume for Session 3. The ipsilesional frontal and lateral temporal lobes show similar downward trends, but do not reach statistical significance. Interestingly, the ipsilesional medial temporal and occipital lobes show patterns of volume increase, though not significant.

**Figure 4.**
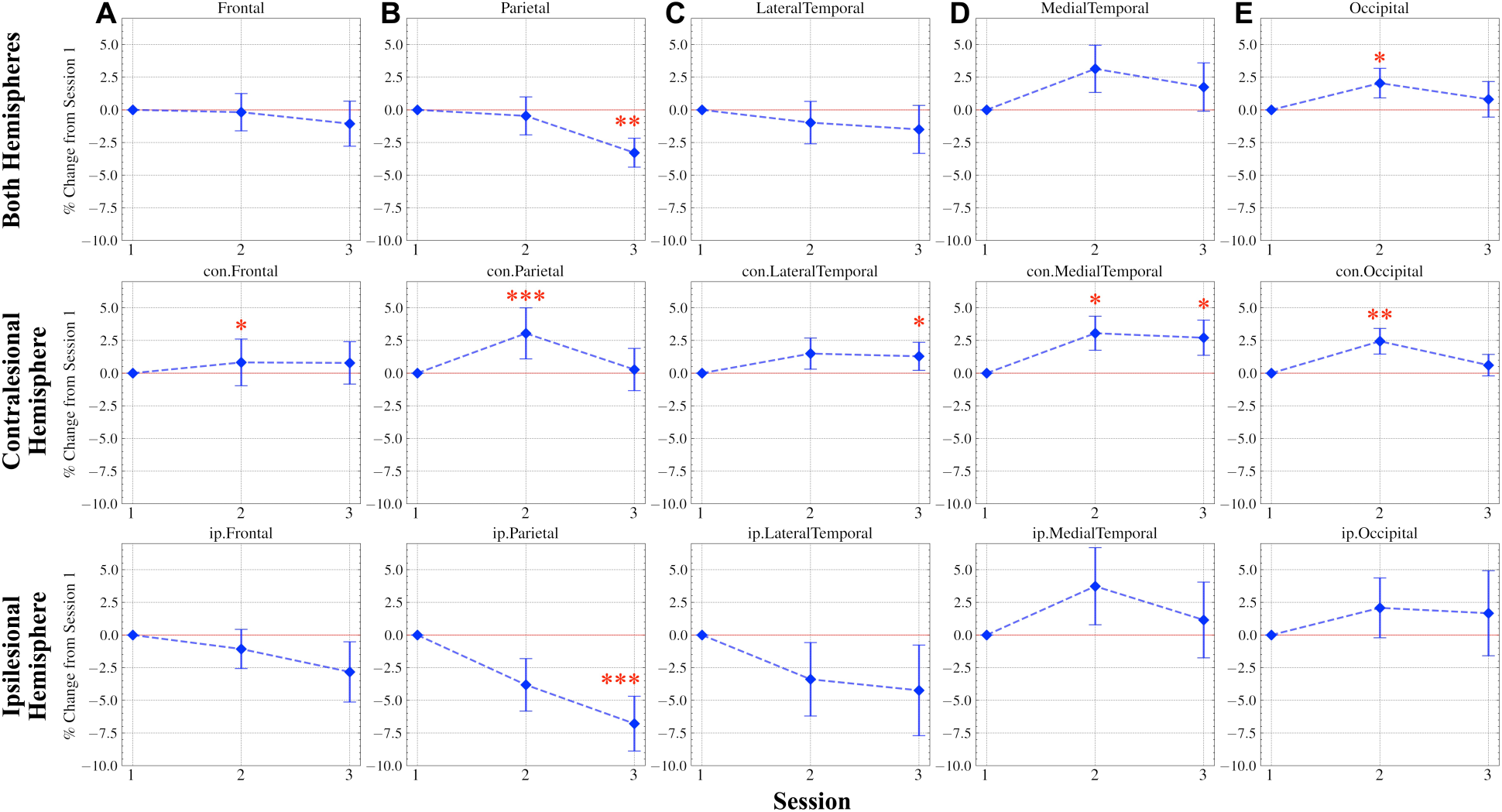
Ipsilesional reduction of brain volume in cortical GM across lobes versus widespread contralesional expansion of cortical GM across lobes. (**A**) Frontal, (**B**) Parietal, (**C**) Lateral Temporal, (**D**) Medial Temporal, and (**E**) Occipital lobes are grouped by (*Top*) Both Hemispheres, (*Middle*) Contralesional Hemisphere, and (*Bottom*) Ipsilesional Hemisphere. * *p* < 0.05, ** *p* < 0.01, *** *p* < 0.001.

Again, by examining the parietal lobe, which displays robust differences both contralesionally and ipsilesionally, we observe that expansion of the contralesional parietal lobe reaches a peak in Session 2 followed by a return to baseline, and shrinkage of the ipsilesional parietal lobe continues downward in Session 3.

Our normalized analysis (Figure S5, Table S5B) corroborates these results, displaying strong contralesional involvement particularly in Session 2 of the frontal [*U* = 37.50, *p* < 0.001, *d* = –0.64] and parietal [*U* = 30, *p* < 0.001, *d* = –0.73] lobes as well as Session 3 of the parietal [*U* = 52.50, *p* = 0.031, *d* = –0.32] and occipital [*U* = 45, *p* = 0.003, *d* = –1.27] lobes. The increasing trends of the contralesional hemisphere are evident across all lobes. The ipsilesional hemisphere shows decreasing trends for all lobes excluding the medial temporal and occipital lobes, as observed unnormalized, with only the parietal lobe reaching significance in its decrease for Session 3 [*U* = 135, *p* = 0.017, *d* = 1.01].

#### Lobar Cortical Thickness

The time evolution of lobar cortical thicknesses (Figure 5, Table S6) is similar to the trajectory of the lobar volumes (Figure 4). However, the lobes with significant changes—the frontal, parietal, and occipital—exhibit a more paired consistency between the contralesional and ipsilesional hemispheres for cortical thickness. Again, contralesionally, we observe peak increase at Session 2. Ipsilesionally, we observe downward trends continuing to Session 3.

**Figure 5.**
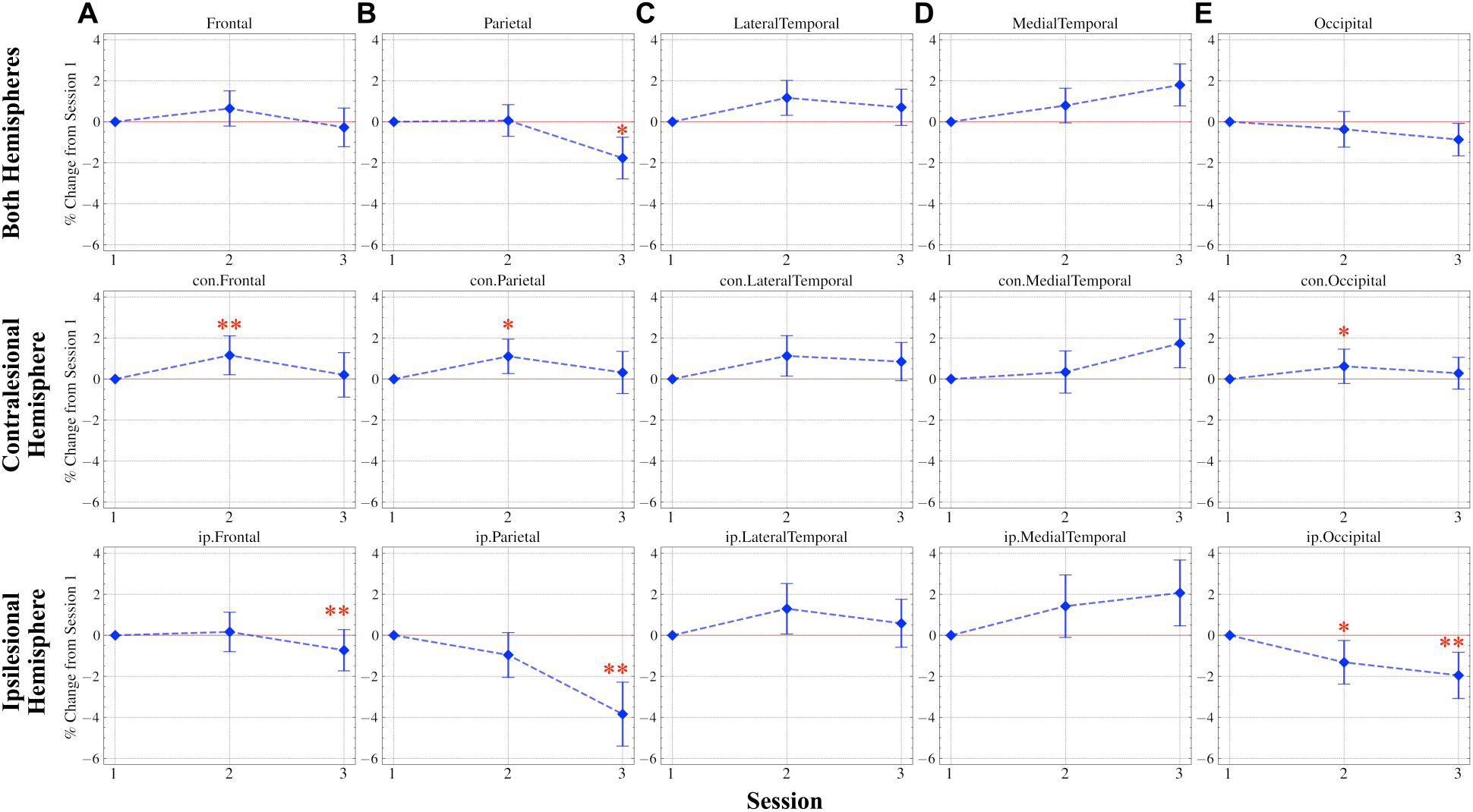
Widespread ipsilesional decrease versus contralesional increase of lobar cortical thickness. (**A**) Frontal, (**B**) Parietal, (**C**) Lateral Temporal, (**D**) Medial Temporal, and (**E**) Occipital lobes are grouped by (*Top*) Both Hemispheres, (*Middle*) Contralesional Hemisphere, and (*Bottom*) Ipsilesional Hemisphere. * *p* < 0.05, ** *p* < 0.01, *** *p* < 0.001.

Interestingly, the lateral and medial temporal lobes don’t exhibit robust changes but positive trends for both the contralesional and ipsilesional hemispheres.

#### Corpus Callosum and Cerebrospinal Fluid Volumes

The corpus callosum (CC) displays significant widespread atrophy, except for its anterior segment (Figure S6A, Table S7A). The mid anterior to posterior CC atrophy is consistent with Wallerian degeneration of commissural fibers affected by the primarily posterior frontal, parietal and temporal lesions. The mid posterior CC, which interfaces between homologous prefrontal, parietal, and posterior temporal cortical areas, exhibits the most notable atrophy and corresponds to the salient parietal lobe volume and cortical thickness changes depicted in Figures 4 and 5. Again, Session 3 exhibits the greatest volume reductions. Our normalized analysis corroborates these results, displaying widespread atrophy (Fig. S6B, Table S7B). Similarly, the mid posterior CC displays the most significant volume decreases at Session 2 [*U* = 180, *p* = 0.003, *d* = 0.67] and Session 3 [*U* = 150, *p* = 0.001, *d* = 1.11]. The Mid Anterior CC also shows significant decrease at Session 3 [*U* = 135, *p* = 0.017, *d* = 0.70].

Cerebrospinal fluid (CSF) volume was shown to increase over sessions for both non-normalized (Fig. S7A, Table S8A) and normalized (Fig. S7B, Table S8B) analyses. For both analyses, significance is reached at Session 3 [non-normalized: *U* = 100, *p* = 0.037, *d* = –0.99; normalized: *U* = 45, *p* = 0.017, *d* = –1.07]. For the normalized analysis, there additionally exists a significant difference between Sessions 2 and 3 [*U* = 47, *p* = 0.038, *d* = –0.68].

#### Correlations in Volume and Cortical Thickness Changes for Homologous Regions and Lobes

Homologous brain regions showed correlated changes in cortical volume and in cortical thickness, such that positive changes in ipsilesional regions tend to be associated with positive changes in contralesional homologous regions and negative changes in ipsilesional regions tend to be associated with negative changes in contralesional homologous regions (Figure 6, Figure S8A, Table S9A). This mirroring pattern exists for both Sessions 2 and 3 and is observed more strongly for cortical thickness. Additionally, the mirroring extends to lobes (Figure 6, Figure S8A, Table S9A). The correlations in changes between homologous lobes and regions display a unique converging pattern of brain recovery between hemispheres as opposed to the diverging trend observed more globally. The normalized analysis of region and lobe volumes (Figure S8B, Table S9B) supports the results observed here, apparent in the trends shown in the scatter plots. However, the exclusion of participants with less eTIV consistency may have contributed to weaker correlations in the normalized analysis.

**Figure 6.**
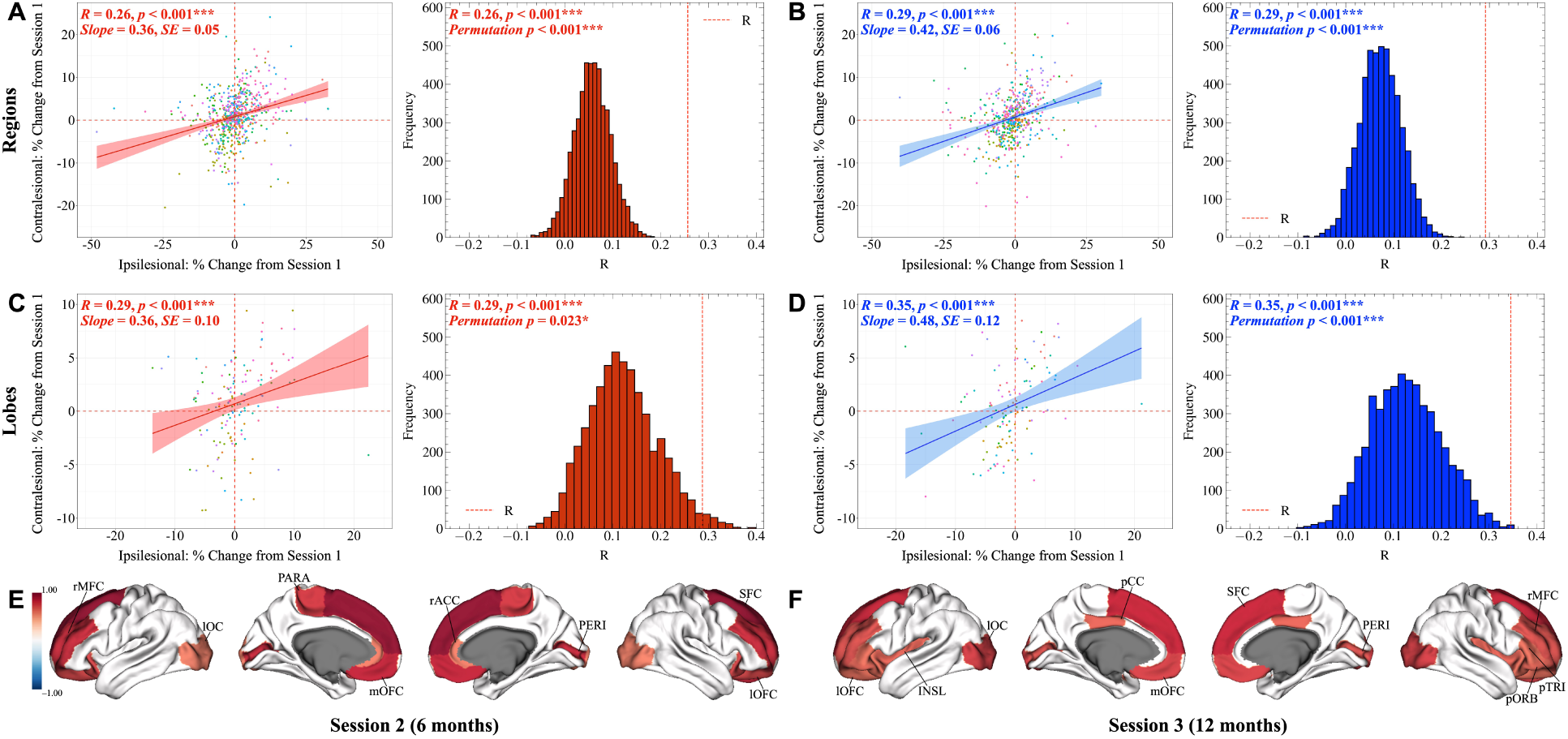
Contralesional brain regions mirror the cortical thickness changes of their homologous counterparts in the ipsilesional hemisphere in children during the first year after ICH. (**A** to **D**) Red shading indicates Session 2, and blue shading indicates Session 3. Scatter plots with the regression line and 95% confidence interval are shown alongside corresponding permutation plots. (**E** to **F**) Regions with robust, significant homologous correlations. * *p* < 0.05, ** *p* < 0.01, *** *p* < 0.001. rMFC, rostral middle frontal cortex; lOC, lateral occipital cortex; PARA, paracentral; mOFC, medial orbitofrontal cortex; rACC, rostral anterior cingulate cortex; PERI, pericalcarine; SFC, superior frontal cortex; lOFC, lateral orbitofrontal cortex; INSL, insula; pCC, posterior cingulate cortex; pORB, pars orbitalis; pTRI, pars triangularis.

We conducted further analyses to examine the specific lobes and regions with the strongest homologous correlations (Fig. 6, Fig. S8A). Correlations in cortical thickness changes between the homologous superior frontal, medial orbitofrontal, pericalcarine, rostral middle frontal, paracentral, lateral orbitofrontal, rostral anterior cingulate, and lateral occipital regions were strongest at Session 2 in comparison to Session 1, in descending order of strength (Fig. S9, Table S10). These regions mainly highlight the frontal lobe, occipital lobe, and cingulate cortex. For Session 3, strong correlations in cortical thickness changes occurred between homologous superior frontal, lateral occipital, medial orbitofrontal, rostral middle frontal, pericalcarine, posterior cingulate, insula, pars orbitalis, lateral orbitofrontal, and pars triangularis regions (in descending order of strength), largely reflecting the lobes and regions identified in Session 2 (Fig. S10, Table S10). For volume, significant, strong correlations existed between the homologous lateral orbitofrontal and paracentral regions in Session 2 (in descending order of strength) (Fig. S8A), but none were significant for Session 3, following FDR BH corrections, which were employed for all correlational analyses in this section. Overall, most correlations in macrostructural changes manifest in terms of cortical thickness, with notable frontal, occipital, and cingulate involvement present at both time points.

#### Correlations in Cortical Thickness Changes for Homologous and Non-Homologous Region Pairs Stratified by Atlases

We extended our analyses beyond homologous connections to include all non-homologous pairs of DKT regions and visualized these patterns using circular connectograms and glass-brain maps for Session 2 (Fig. 7) and Session 3 (Fig. 8). Across both sessions, contralesional cortical alterations predominated over ipsilesional changes, and these were primarily expressed as robust, positive correlations. The most frequent alterations in connections involved frontal–frontal and frontal–parietal regions, with additional contributions from frontal–temporal interactions. The strongest significant edges are visualized in plots and summarized in tables, which are provided in the Supplementary Material for Sessions 2 (Fig. S11, Table S11) and 3 (Fig. S12, Table S11).

**Figure 7.**
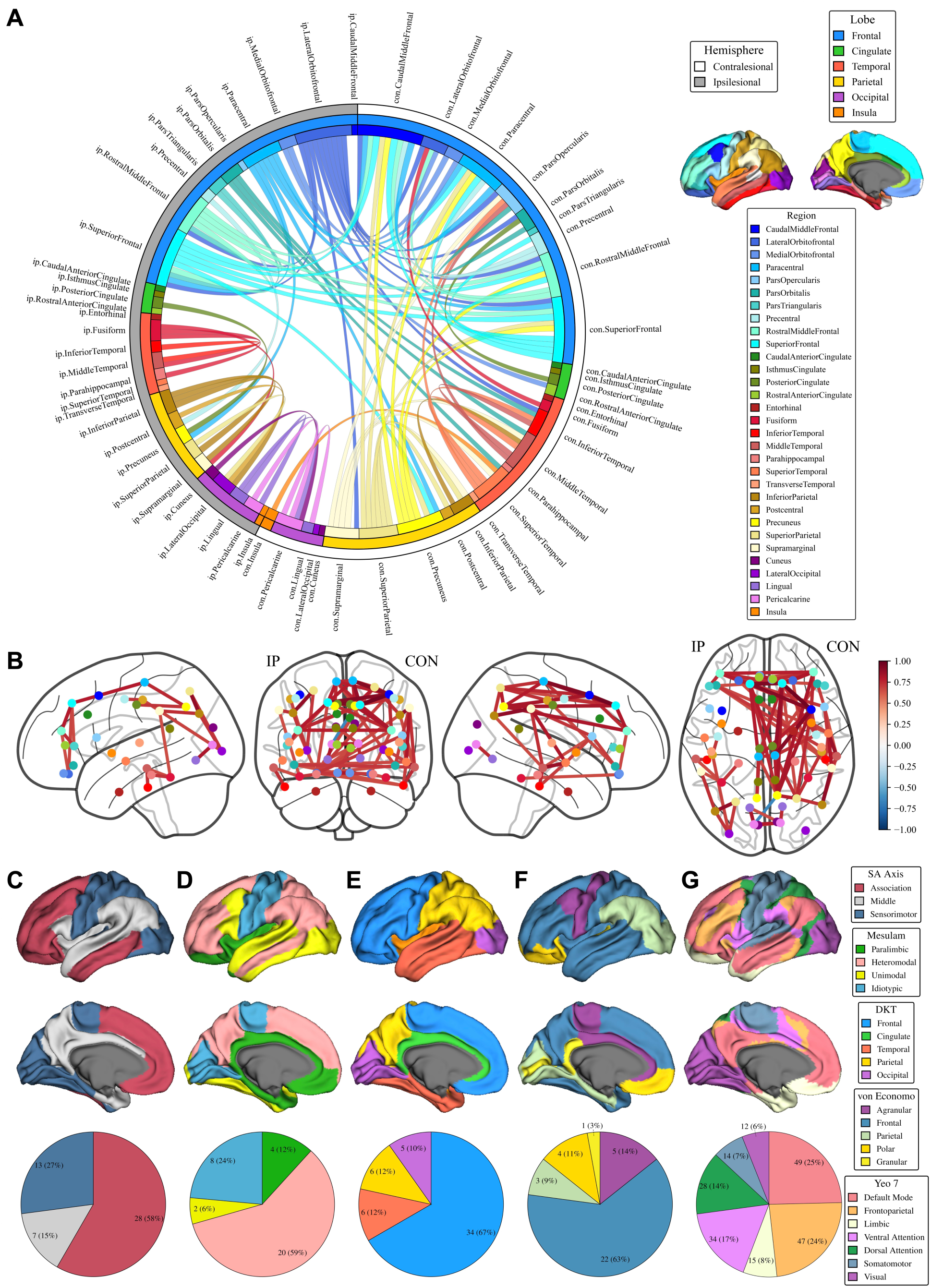
Homologous and non-homologous correlations of regional cortical thickness reveal pronounced contralesional involvement, with alterations primarily in association, heteromodal, and frontal cortices, as well as default mode and frontoparietal networks, at 6 months from baseline. (**A**) Circular connectogram with hemisphere indicated in the outermost ring, lobe in the middle ring, and region in the innermost ring. Length of each area’s ring link corresponds to its number of significant connections, or edges. Plotted according to Desikan-Killiany-Tourville (DKT) regions defined in the legend and brain plot. (**B**) Glass-brain map with nodes as DKT regions using the same color scheme as the DKT legend. (**C** to **G**) Brain plots and pie charts indicating the proportion of significant edges across classes of the (**C**) sensorimotor–association (SA) axis, (**D**) Mesulam, (**E**) Desikan-Killiany-Tourville, (**F**) von Economo-Koskinas, and (**G**) Yeo 7-Network atlases. For each class, the number of edges and the proportion (% in parentheses) are provided. CON, contralesional; IP, ipsilesional.

**Figure 8.**
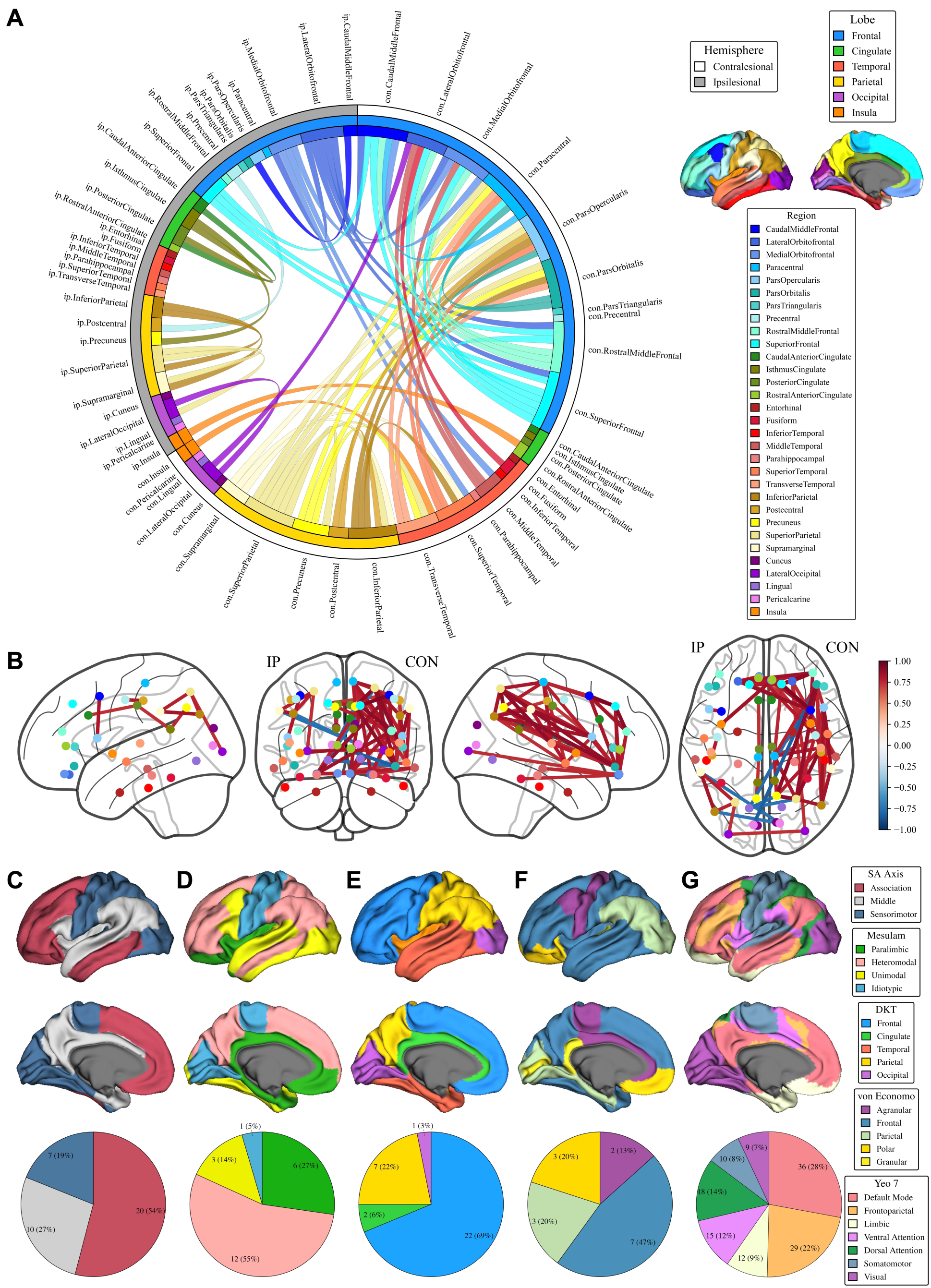
Homologous and non-homologous correlations of regional cortical thickness reveal pronounced contralesional involvement, with alterations primarily in association, heteromodal, and frontal cortices, as well as default mode and frontoparietal networks, at 12 months from baseline. (**A**) Circular connectogram with hemisphere indicated in the outermost ring, lobe in the middle ring, and region in the innermost ring. Length of each area’s ring link corresponds to its number of significant connections, or edges. Plotted according to Desikan-Killiany-Tourville (DKT) regions defined in the legend and brain plot. (**B**) Glass-brain map with nodes as DKT regions using the same color scheme as the DKT legend. (**C** to **G**) Brain plots and pie charts indicating the proportion of significant edges across classes of the (**C**) sensorimotor–association (SA) axis, (**D**) Mesulam, (**E**) Desikan-Killiany-Tourville, (**F**) von Economo-Koskinas, and (**G**) Yeo 7-Network atlases. For each class, the number of edges and the proportion (% in parentheses) are provided. CON, contralesional; IP, ipsilesional.

To contextualize these findings, we stratified the significant edges using multiple atlas frameworks, including the SA, Mesulam, DKT, von Economo-Koskinas, and Yeo 7-Network fMRI atlases (Fig. 7 & Fig. 8). This stratification reinforced the patterns observed in the circular connectogram and glass-brain map results. Most significant connectogram edges localized to the association pole of the SA axis, heteromodal cortex in Mesulam’s hierarchy of laminar differentiation, and frontal cortex among the von Economo-Koskinas cytoarchitectural types. At the intrinsic connectivity network level, the default mode and frontoparietal networks accounted for the largest proportion of significant edges, followed by the ventral attention and dorsal attention networks. This spatial distribution was consistently observed across both Sessions 2 and 3.

## DISCUSSION

Our longitudinal neuroimaging examination of children following ICH advances the understanding of morphological alterations that occur during the first year of recovery. While prior studies of cortical plasticity due to stroke have primarily focused on ischemic stroke in adults, our investigation of ICH in children offers comparative, exploratory insights into a crucial and understudied population. Moreover, we employ novel analytic approaches to this population, including a structural covariance framework, that captures coordinated changes within and between hemispheres, and a hierarchical categorization of brain regions, including categorization into fMRI networks, thereby allowing for a more nuanced understanding of the systematic brain changes underlying recovery from pediatric ICH. Our key observations include: (1) opposing macrostructural changes—contralesional increases and ipsilesional decreases in cortical volume and thickness—of the left and right cerebral hemispheres; (2) mirrored changes between homologous lobes and regions across cerebral hemispheres, particularly of cortical thickness; (3) morphological alterations that predominantly involve higher-order cortex of association, heteromodal, and frontal classes, especially within the default mode, frontoparietal, ventral attention, and dorsal attention networks; (4) topographic callosal Wallerian degeneration reflecting the lesion location; and (5) differential temporal dynamics of cortical thickness changes characterized by contralesional increases peaking at 6 months versus more sustained ipsilesional decreases that intensify by one year post-ICH.

As hypothesized, contralesional hemispheric involvement emerged as a key feature of recovery, consistent with extensive prior stroke literature^23–27^. Notably, contralesional involvement was marked by increases in volume and cortical thickness, in contrast to ipsilesional decreases, especially in perilesional areas. While atrophy of the ipsilesional hemisphere is expected and has been observed even 18 to 36 months post-stroke^57^, we observe that the two hemispheres display opposing trajectories over time in volume and cortical thickness, including across homologous lobe pairs.

While the aforementioned hemispheric and lobar alterations suggest diverging neuroplastic mechanisms between the cerebral hemispheres, at a more granular level, regional analyses revealed that changes in volume and cortical thickness jointly correspond, mainly positively, between homologous brain regions, as observed in previous studies^27,57–61^. It is important to note that decreases in cortical volume and thickness also occur in the intact contralesional hemisphere, along with bilateral decreases^27,58,61^, as noted in prior studies. The general pattern of coupled alterations was especially robust for cortical thickness in homologous regions of the frontal and occipital lobes, as well as cingulate cortex. Based on prior literature, this pattern would indicate that morphological changes of the brain during recovery from ICH reflect a pattern of structural covariance—similarities in morphological properties between structures, especially homologous pairs, across the two hemispheres. Structural covariance has long been observed both cross-sectionally and in the coordinated trajectories of cortical maturation^62,63^. This same tendency may also extend to longitudinal morphometric alterations that follow localized brain injury, manifesting at the level of regional change.

Overall, this pattern of homologous reactivity suggests that recovery mechanisms aim to maintain bilateral symmetry between hemispheres through compensatory recruitment. This tendency toward balance and symmetry reinforces the brain’s inherently bilateral architecture, evident in structural covariance networks^17,18^ and resting-state fMRI networks^19,20^ (e.g., the visual network encompasses both left and right visual cortices), suggesting that brain function, neural connectivity, and recovery mechanisms ultimately rely on, as well as reflect, homologously paired wiring^21,22^. Post-ICH cortical plasticity conforms to homologous disparity, but it is not yet evident whether this is beneficial or detrimental to recovery and functioning, as evidence is provided for both cases^24,64^.

We observe that the majority of post-ICH alterations, whether homologous or non-homologous, cluster within association cortex, particularly regions of heteromodal laminar differentiation and frontal cytoarchitecture. The participation of the default mode, frontoparietal, ventral attention, and dorsal attention networks highlights a common locus of cortical reorganization. These higher-order systems are critical for integrating multimodal information and orchestrating flexible introspective as well as goal-directed behavior^36,65^. Their preferential involvement suggests that the impact of focal vascular injury extends well beyond primary sensorimotor territories to disrupt the brain’s integrative hubs, as previously observed in regions such as the orbitofrontal cortex and middle frontal gyri^66^. Furthermore, neuroimaging studies of ischemic stroke patients have shown that neuroplasticity extends far beyond the lesion and its contralateral counterpart, engaging distant cortical^28,29^ and subcortical^30^ regions not typically involved in the impaired functions^67,68^. Given the dense interconnectivity and high metabolic demands of these brain networks, they may be particularly susceptible to focal lesions^69^, consistent with theories of diaschisis^70,71^ and network vulnerability^72^.

At the same time, these late-developing regions are also thought to possess heightened plasticity^73–75^. Their malleability may underlie not only vulnerability but also opportunities for reorganization. Strikingly, a predominance of robust, significant region-to-region homologous and non-homologous correlations emerge within the contralesional hemisphere, indicating that much of the reorganization is occurring in uninjured tissue. This suggests that compensatory processes are being actively sourced from regions where structural integrity remains intact.

Furthermore, increases in contralesional volume and cortical thickness have been associated with greater motor recovery and improved clinical outcomes after ischemic stroke^60,76^, particularly when involving increased GM volume in cognitive regions^58^. Conversely, decreases in GM volume and cortical thickness have been associated with poorer outcomes. Altogether, the integrative and flexible nature of higher-order networks may also provide a substrate for recovery that draws particularly on the contralesional hemisphere to supply neural resources and facilitate cortical reorganization.

The structural integrity of the corpus callosum, crucial for interhemispheric communication^77^, shows notable decline, especially in the mid posterior segment, which is known to bridge between the prefrontal, parietal, and temporal lobes, as well as the central segment (body), which connects between homologous motor and somatosensory areas. The locations of callosal atrophy correspond topographically with commissural WM fiber bundles projecting to locations where our lesions predominantly occur, consistent with Wallerian degeneration.

Broadly, we notice that changes from baseline persist in Session 3, one-year post-injury. Increases in volume and cortical thickness emerge contralesionally at Session 2 but subsequently dip back toward baseline by Session 3. In contrast, ipsilesional atrophy not only appears at Session 2 but also continues to worsen into Session 3. Such temporal dynamics—marked by early, transient contralesional increases and more prolonged ipsilesional decreases—suggest temporally dependent hemispheric divergences in the recovery process. Specifically, presumably compensatory responses appear to occur earlier in time and contralesionally, whereas atrophy follows a more extended trajectory ipsilesionally. In parallel, CSF volume increases steadily across sessions, consistent with ongoing post-ICH global atrophy also involving subcortical GM and WM. While existing literature often suggests that most post-stroke changes occur within 3–6 months, these studies seldom explore the sustainability of these changes over longer periods^78,79^. Our findings demonstrate that cortical alterations not only persist^57^ but also diverge across hemispheres, emphasizing the asymmetric time scales of neural compensation and degeneration.

### Limitations and Future Directions

We acknowledge several limitations and uncertainties that provide directions for further investigation. Firstly, the observed predominance of left hemispheric lesions, particularly within the temporoparietal region, raises questions about potential biological tendencies that are related to pediatric ICH or the influence of sampling bias. Secondly, the neurobiological mechanisms underlying changes in brain macrostructure remain elusive and may require advanced imaging techniques, such as microstructural imaging, to decipher. Thirdly, the generalizability of bilateral symmetry in macrostructural changes to other unilateral neurological conditions remains unclear but holds implications for recovery and rehabilitation. Fourthly, patients undergoing complex surgical interventions for ICH, such as coil embolization, clipping, and cerebral shunt placement, often present with images that are not analyzable due to metallic artifacts. This limitation, in addition to patient attrition over one-year follow-up, restricts our analysis, particularly for Session 3. Lastly, the sample size of outcome data was too small to investigate the clinical and cognitive correlates of the observed cortical neuroplasticity. However, future analyses incorporating the extended clinical outcomes of participants who do not overlap with our MRI analysis sample, along with consideration of neurological recovery, may provide further insights into the functional presentation of pediatric ICH patients during recovery.

## Conclusions

Our study reveals that pediatric post-ICH recovery involves alterations in brain morphology that extend beyond the expected perilesional area and even the ipsilesional hemisphere to involve the contralesional hemisphere. Notably, increased volume and cortical thickness are more prominent in the cortex of the contralesional hemisphere during an earlier post-injury period, whereas cortical, subcortical, and white matter atrophy are predominantly observed in the ipsilesional hemisphere over a longer time scale. At a more granular level, cortical reorganization is consistent across homologous regions of the two hemispheres, reflecting a pattern of joint adaptive neuroplasticity versus joint neurodegeneration. In general, most cortical alterations following pediatric ICH involve association, heteromodal, and frontal cortices of the contralesional hemisphere, especially within the default mode, frontoparietal, ventral attention, and dorsal attention networks. This pattern highlights both the vulnerability and inherent plasticity of higher-order systems that support cognition, attention, and executive function, and suggests that these same networks—drawing particularly on the intact contralesional hemisphere—may serve as substrates of resilience during recovery. Lastly, the underlying biological mechanisms driving these changes and their specific directionality remain to be elucidated, as well as their clinical, cognitive, and behavioral consequences.

## ARTICLE INFORMATION

Supplemental Material is available.

Abstract presented at the International Stroke Conference, February 7–9, 2024. To be presented at the International Pediatric Stroke Organization Congress, May 27–29, 2026.

## Data Availability

All data produced in the present study are available upon reasonable request.

## Acknowledgments

We especially thank our participants and their families for their generosity and time, granting us the opportunity to learn more about ICH in children and share our findings with the community.

## Abbreviations and Acronyms

AVM: arteriovenous malformation
BH: Benjamini-Hochberg
CC: corpus callosum
CM: cavernous malformation
CSF: cerebrospinal fluid
DKT: Desikan-Killiany-Tourville
eTIV: estimated total intracranial volume
fMRI: functional magnetic resonance imaging
GM: gray matter
ICH: intracerebral hemorrhage
MP-RAGE: magnetization-prepared rapid gradient-echo
MRI: magnetic resonance imaging
SA: sensorimotor–association
T1w: T1-weighted
T2-FLAIR: T2-weighted fluid-attenuated inversion recovery
WM: white matter

