## Supplemental Material for "Cerebral Cortical Reorganization After Intracerebral Hemorrhage in Children"

**This PDF file includes:**

Supplementary Figures 1 to 12
Supplementary Tables 1 to 11

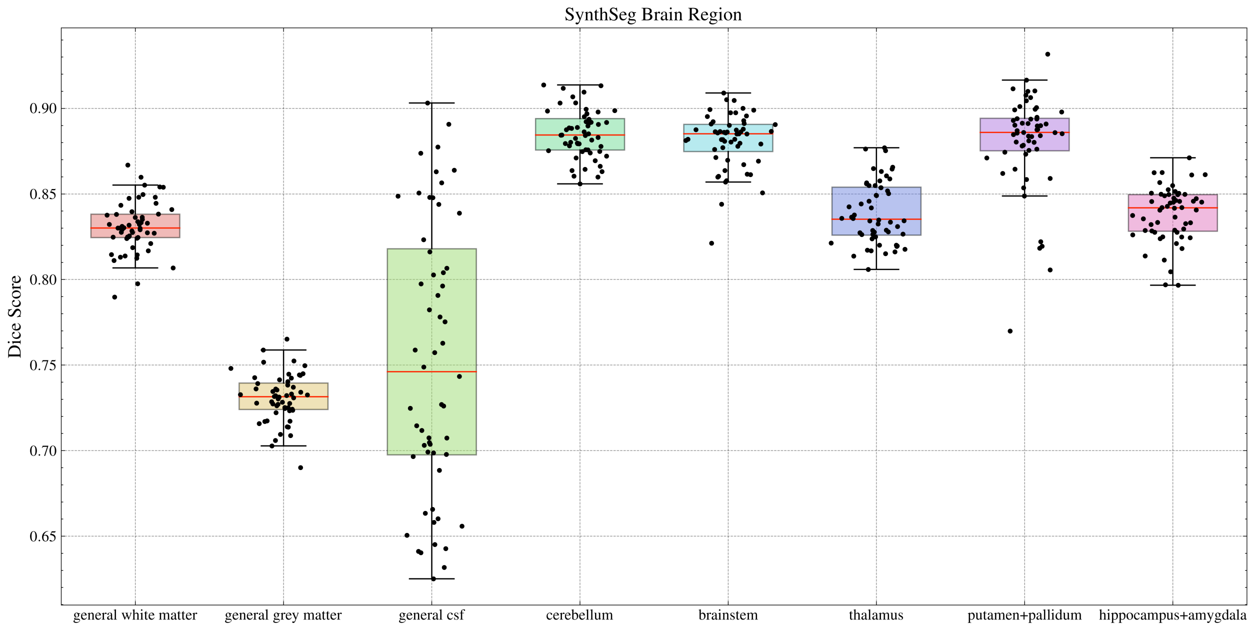


**Figure S1. SynthSeg segmentation performance indicated by dice scores.** SynthSeg provides measures of segmentation performance in Dice scores by testing the predicted segmentation of select regions to a reference ground-truth[1]. CSF, cerebrospinal fluid.

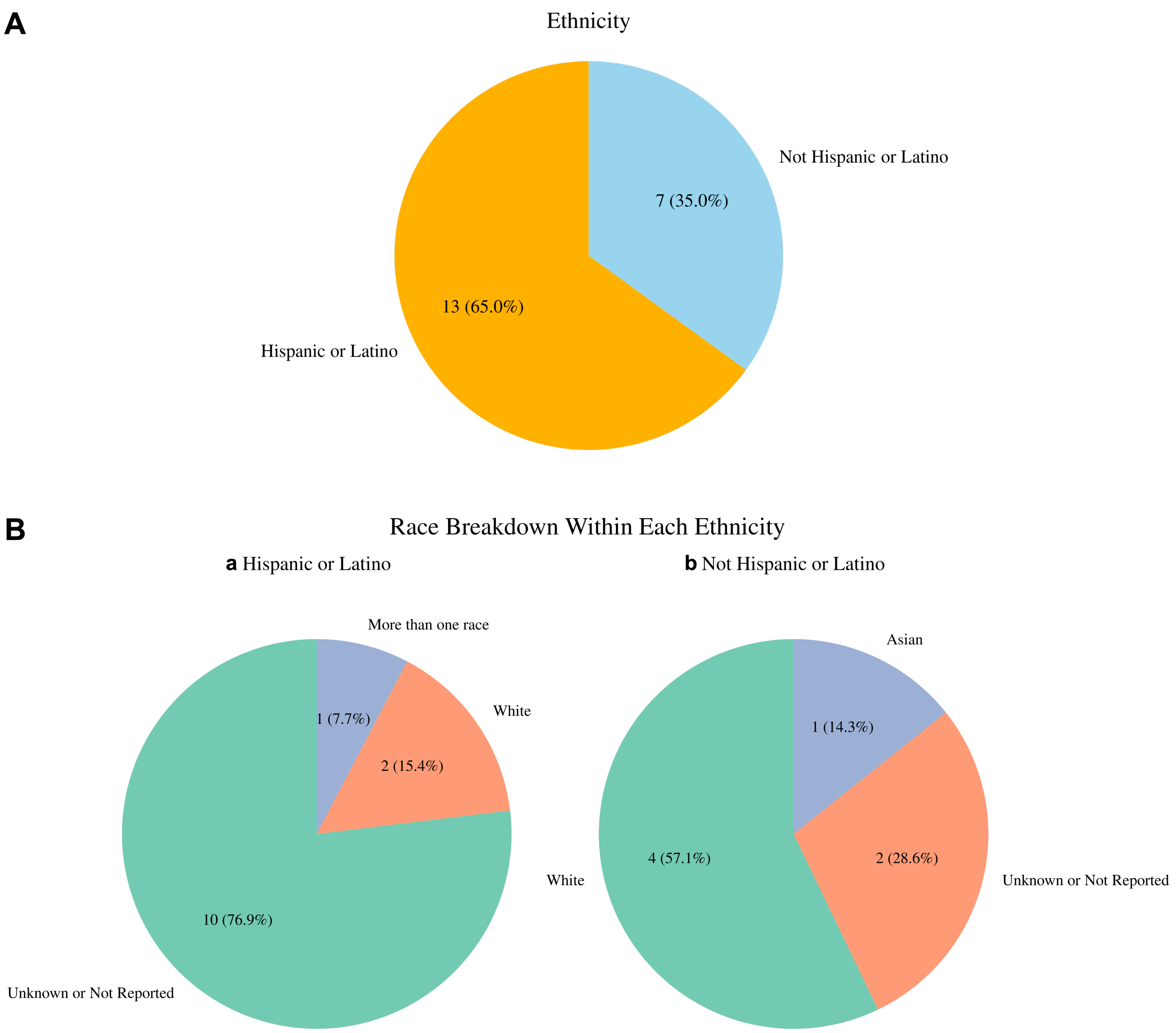


**Figure S2. Ethnic and racial demographics of participants.** (*Top*) General ethnic groupings of Hispanic or Latino (orange) versus Not Hispanic or Latino (blue). (*Bottom*) Breakdown of general ethnic groupings into race. Green indicates largest proportion, followed by red and purple. Participant *n* and their proportions (% in parentheses) are provided.

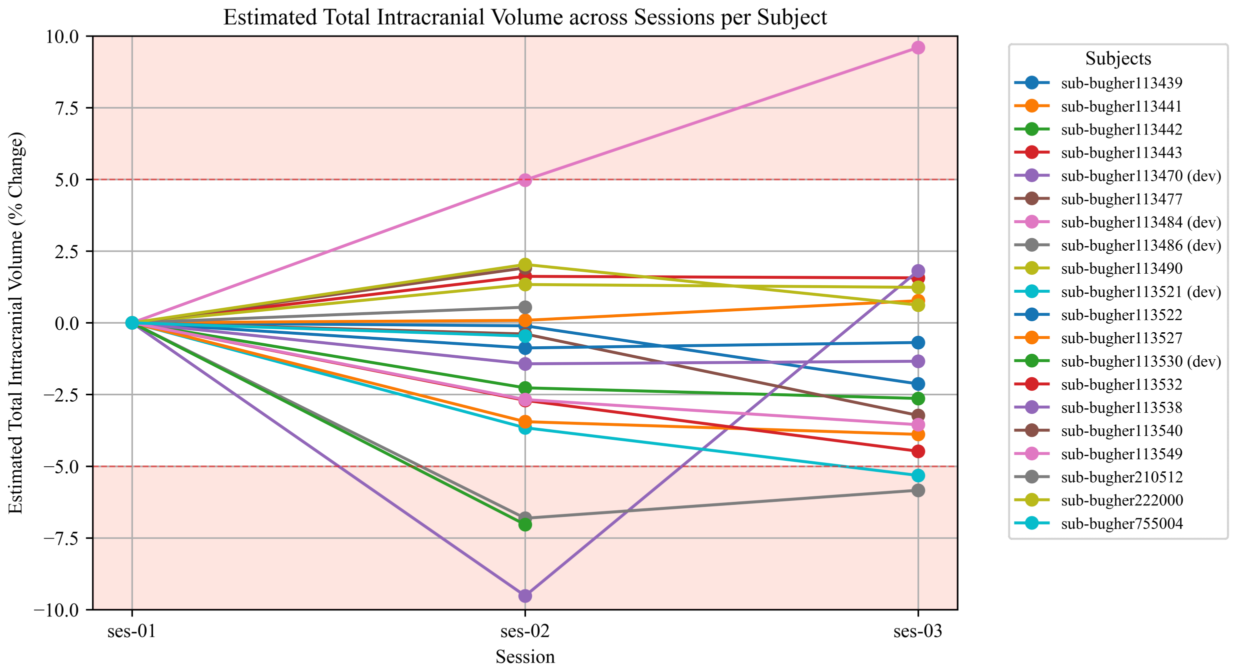


**Figure S3. Estimated total intracranial volume of participants across sessions.** Participants who deviated (dev) over 5 percent from baseline, positively or negatively, are indicated in red shading and in the participant key to the right.

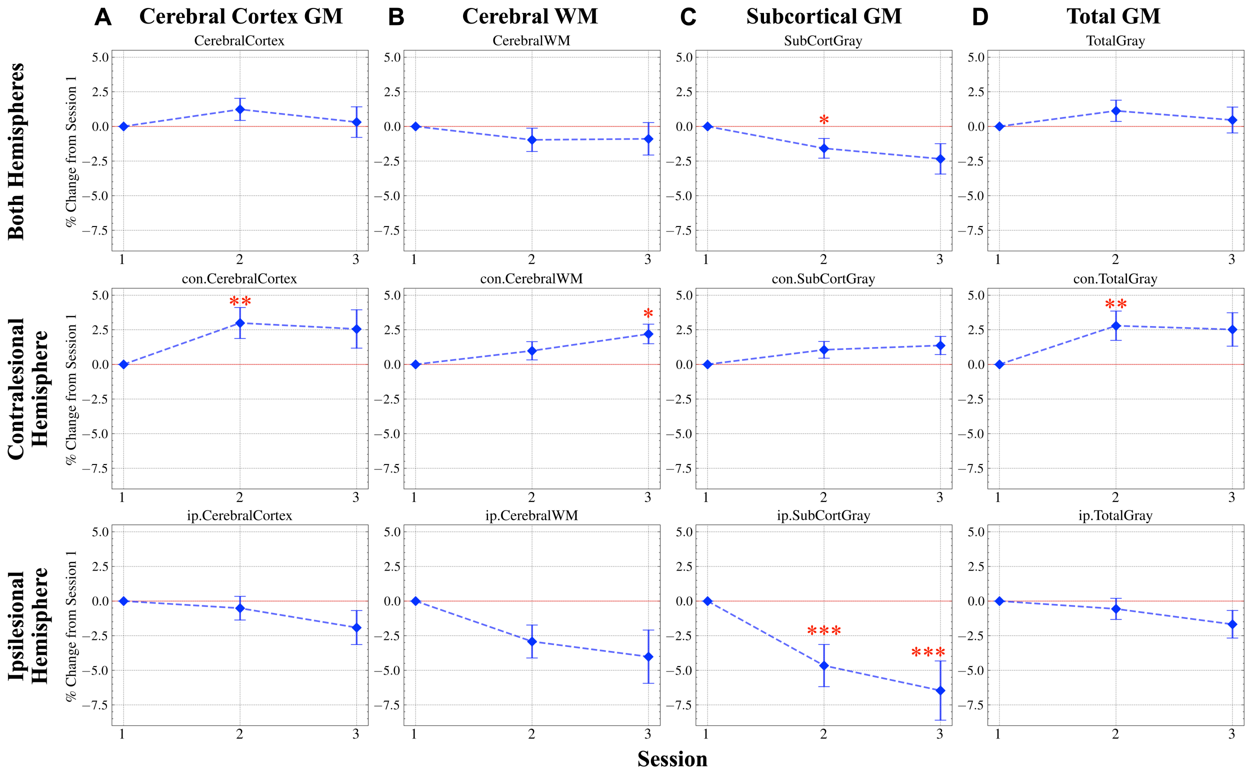


**Figure S4.** ***Volumes normalized by eTIV:*** **ipsilesional subcortical GM neurodegeneration and contralesional cerebral GM and WM neuroplasticity after pediatric ICH.** (**A**) Cerebral Cortex GM, (**B**) Cerebral WM, (**C**) Subcortical GM, and (**D**) Total GM are grouped by (*Top*) Both Hemispheres, (*Middle*) Contralesional Hemisphere, and (*Bottom*) Ipsilesional Hemisphere. Session 2 ($n$ = 15) and Session 3 ($n$ = 12) due to the exclusion of 5 participants with less stable eTIV measurements. * $p < 0.05$, ** $p < 0.01$, *** $p < 0.001$. eTIV, estimated total intracranial volume; GM, gray matter; WM, white matter; ICH, intracerebral hemorrhage.

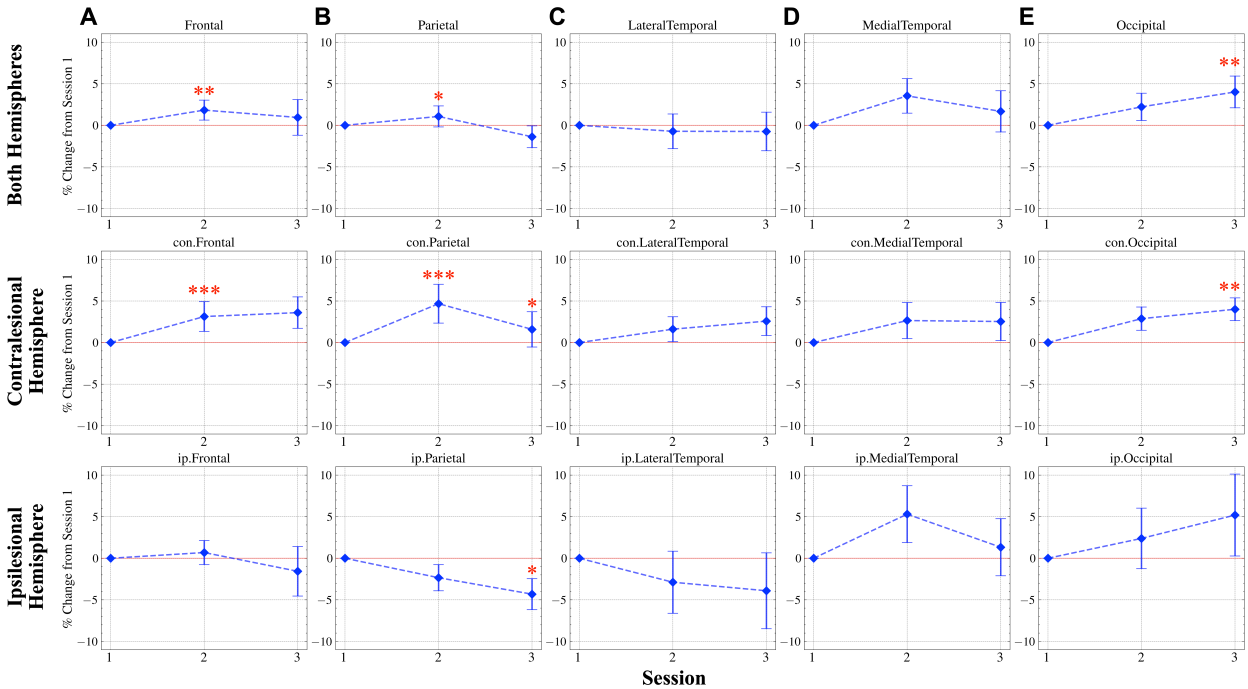


**Figure S5.** ***Volumes normalized by eTIV:*** **ipsilesional neurodegeneration of brain volume in affected lobe GM and widespread contralesional neuroplasticity including homologous parietal, frontal, and occipital lobe GM.** (**A**) Frontal, (**B**) Parietal, (**C**) Lateral Temporal, (**D**) Medial Temporal, and (**E**) Occipital lobes are grouped by (*Top*) Both Hemispheres, (*Middle*) Contralesional Hemisphere, and (*Bottom*) Ipsilesional Hemisphere. Session 2 ($n$ = 15) and Session 3 ($n$ = 12) due to the exclusion of 5 participants with less stable eTIV measurements. * $p < 0.05$, ** $p < 0.01$, *** $p < 0.001$. eTIV, estimated total intracranial volume; GM, gray matter.

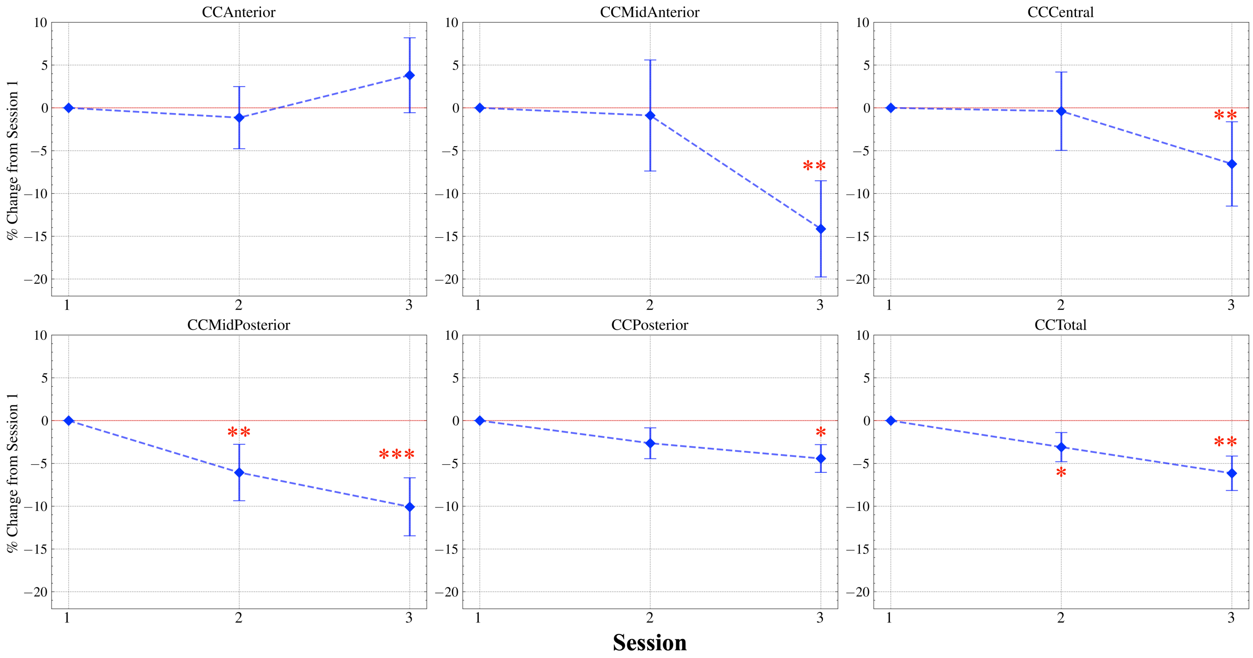


**Figure S6A. Callosal volume degeneration is associated with lesion location, reflecting the topography of posterior frontal, parietal, and temporal lobe inter-hemispheric connectivity.** * $p < 0.05$, ** $p < 0.01$, *** $p < 0.001$. CC, corpus callosum.

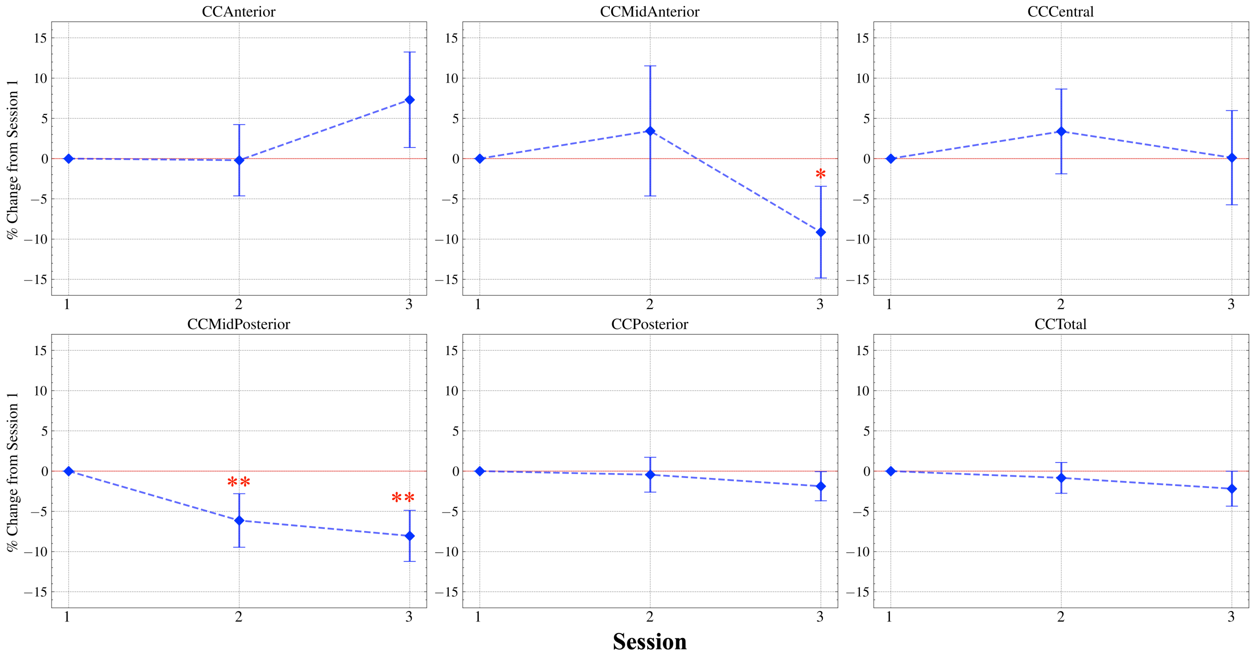


**Figure S6B.** ***Volumes normalized by eTIV:*** **callosal volume degeneration is associated with lesion location, reflecting the topography of posterior frontal, parietal, and temporal lobe inter-hemispheric connectivity.** Session 2 ($n$ = 15) and Session 3 ($n$ = 12) due to the exclusion of 5 participants with less stable eTIV measurements. * $p < 0.05$, ** $p < 0.01$, *** $p < 0.001$. eTIV, estimated total intracranial volume; CC, corpus callosum.

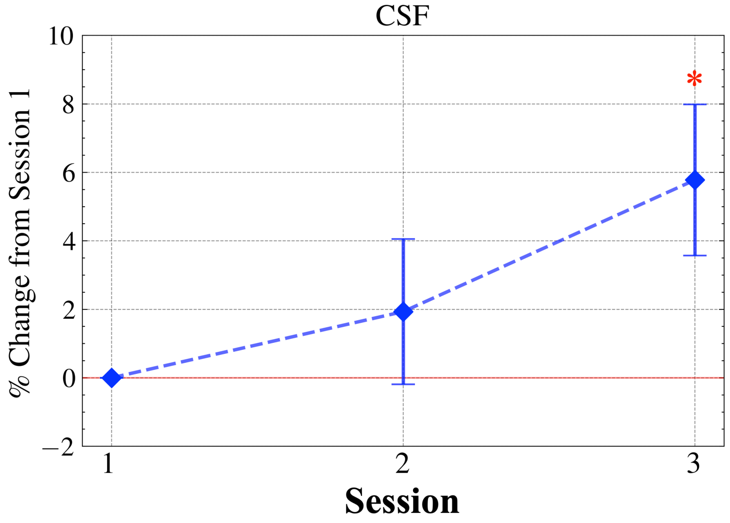


**Figure S7A. CSF volume increase over one year.** * $p < 0.05$, ** $p < 0.01$, *** $p < 0.001$. CSF, cerebrospinal fluid.

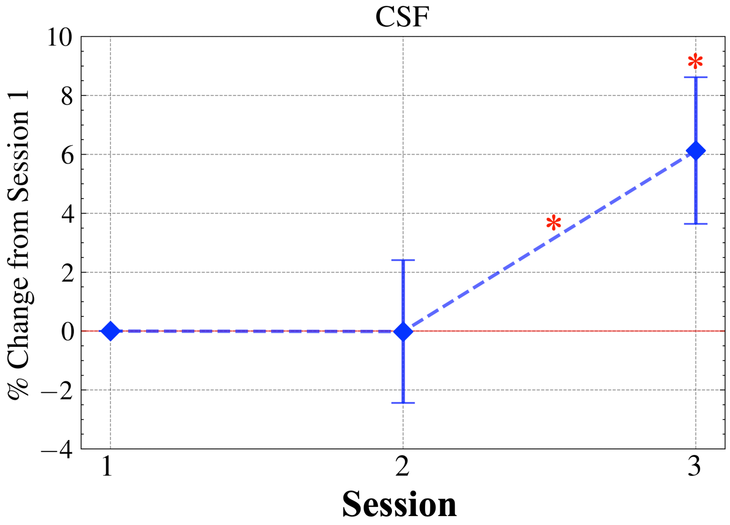


**Figure S7B. *Volume normalized by eTIV:* CSF volume increase over one year.** Session 2 ($n$ = 15) and Session 3 ($n$ = 12) due to the exclusion of 5 participants with less stable eTIV measurements. * $p < 0.05$, ** $p < 0.01$, *** $p < 0.001$. eTIV, estimated total intracranial volume; CSF, cerebrospinal fluid.

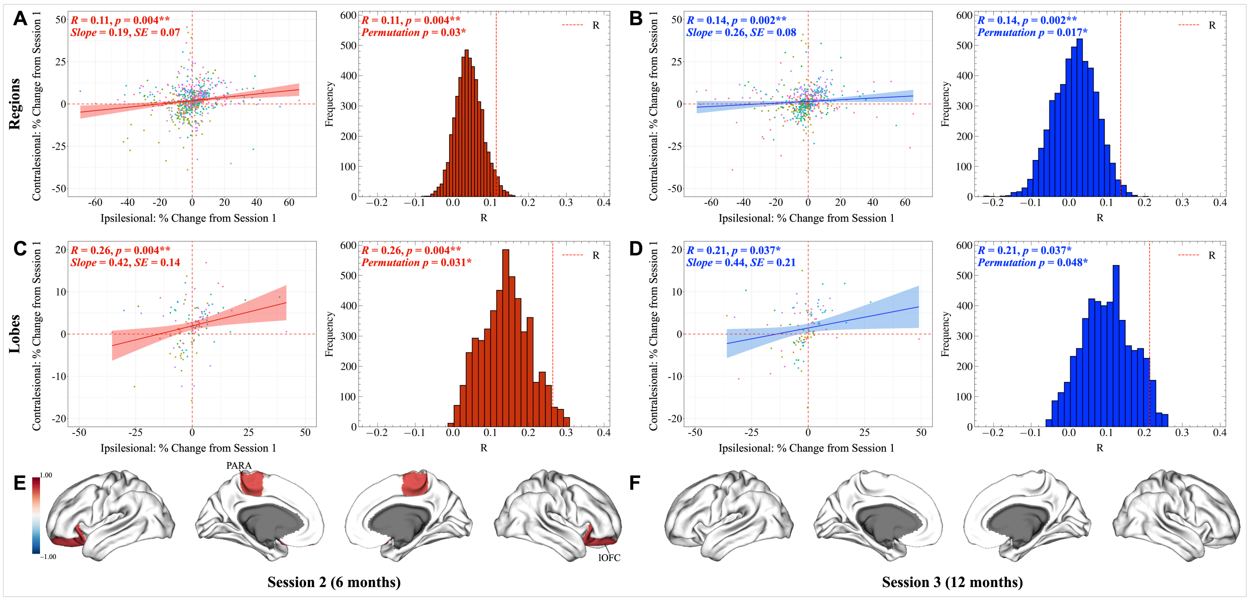


**Figure S8A. Contralesional brain regions and lobes mirror the volume changes of their homologous counterparts in the ipsilesional hemisphere in children during the first year after ICH.** (**A** to **D**) Red shading indicates Session 2 ($n = 20$), and blue shading indicates Session 3 ($n = 16$). Scatter plots with the regression line and 95% confidence interval are shown alongside corresponding permutation plots. (**E** and **F**) Regions with robust, significant homologous correlations following BH FDR corrections. * $p < 0.05$, ** $p < 0.01$, *** $p < 0.001$. ICH, intracerebral hemorrhage; PARA, paracentral; lOFC, lateral orbitofrontal cortex.

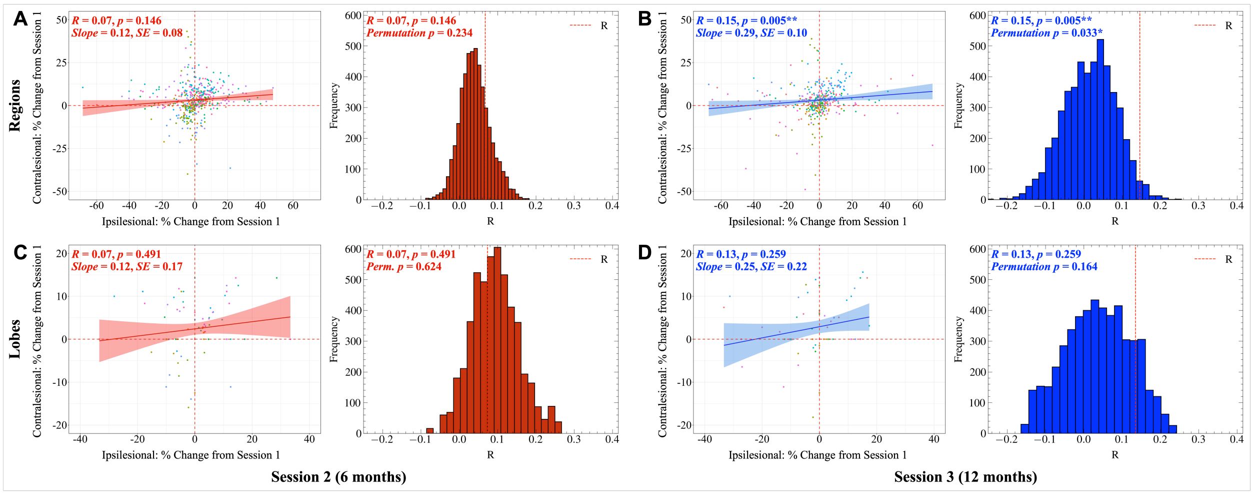


**Figure S8B.** ***Volumes normalized by eTIV:* contralesional brain regions and lobes mirror the volume changes of their homologous counterparts in the ipsilesional hemisphere in children during the first year after ICH.** Red shading indicates Session 2, and blue shading indicates Session 3. Scatter plots with the regression line and 95% confidence interval are shown alongside corresponding permutation plots. Session 2 ($n = 15$) and Session 3 ($n = 12$) due to the exclusion of 5 participants with less stable eTIV measurements. Brain plots are not displayed since no correlations of homologous brain regions were significant following FDR correction using the BH procedure. * $p < 0.05$, ** $p < 0.01$, *** $p < 0.001$. eTIV, estimated total intracranial volume; ICH, intracerebral hemorrhage.

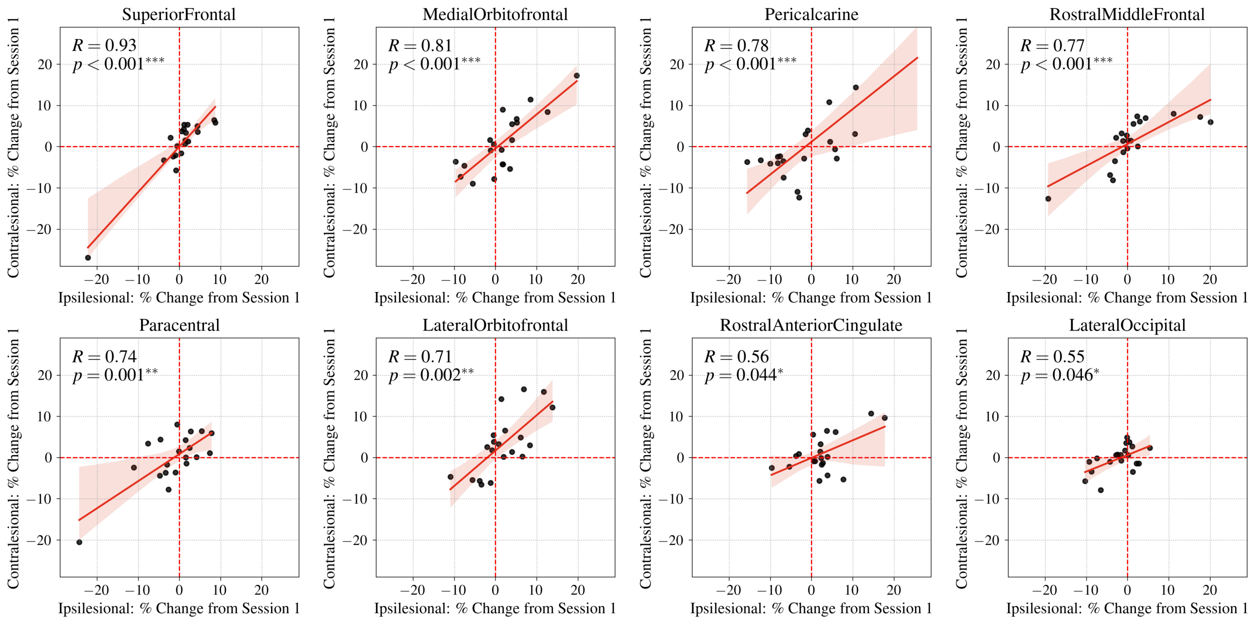


**Figure S9. Regions with robust and significant homologous correlations for cortical thickness at 6 months from baseline.** All significant regions are displayed. Red shading indicates Session 2, 6 months from baseline. Scatter plots with the regression line and 95% confidence interval are shown. The p-values displayed are following FDR correction using the BH procedure. * $p < 0.05$, ** $p < 0.01$, *** $p < 0.001$.

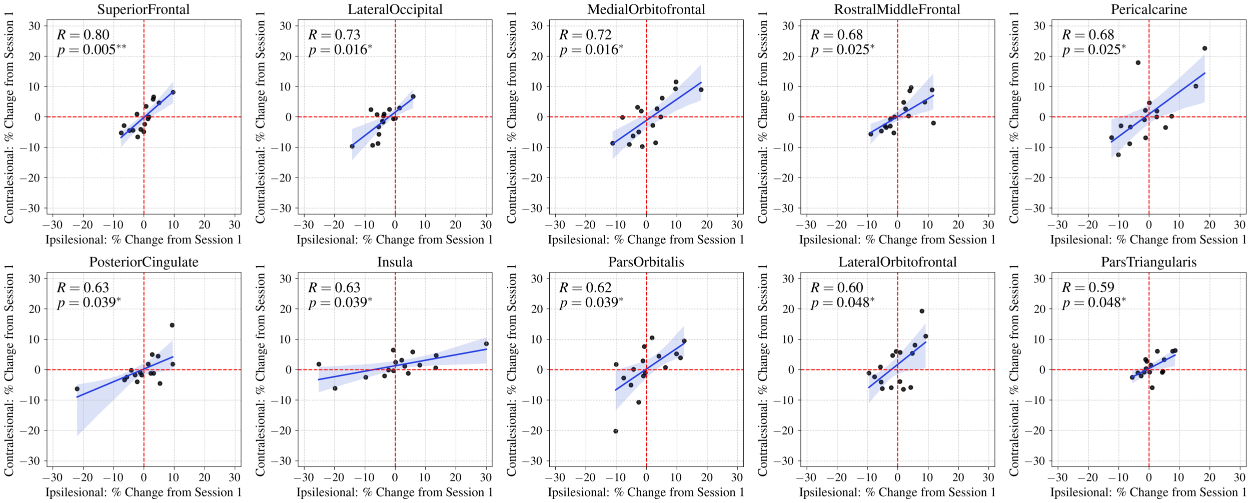


**Figure S10. Regions with robust and significant homologous correlations for cortical thickness at 12 months from baseline.** All significant regions are displayed. Blue shading indicates Session 3, 12 months from baseline. Scatter plots with the regression line and 95% confidence interval are shown. The p-values displayed are following FDR correction using the BH procedure. * $p < 0.05$, ** $p < 0.01$, *** $p < 0.001$.

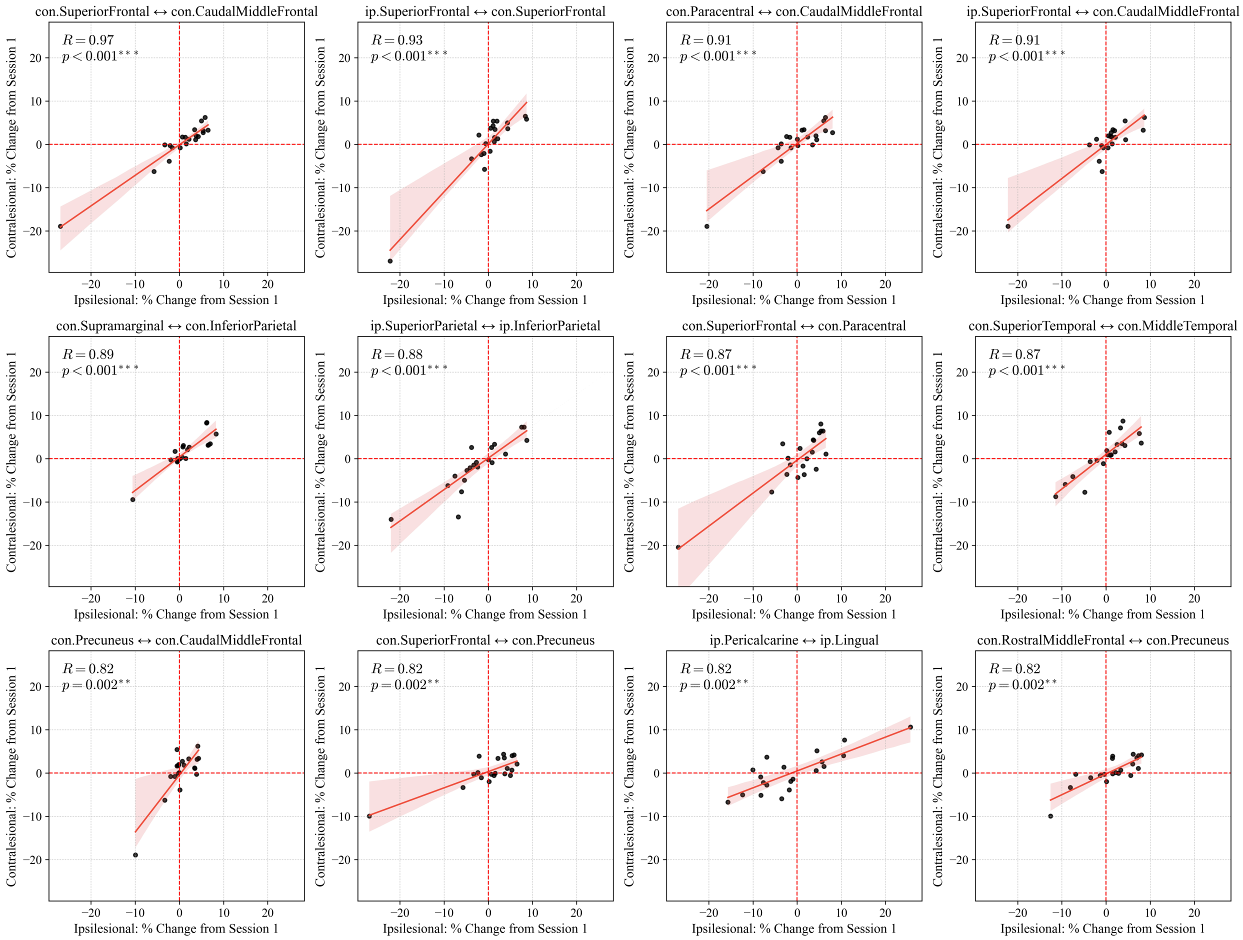


**Figure S11. Non-homologous and homologous region pairs with robust and significant correlations for cortical thickness at 6 months from baseline.** The top 12 significant region pairs (i.e., edges) are displayed. Red shading indicates Session 2, 6 months from baseline. Scatter plots with the regression line and 95% confidence interval are shown. The p-values displayed are following FDR correction using the BH procedure. * $p < 0.05$, ** $p < 0.01$, *** $p < 0.001$. CON, contralesional; IP, ipsilesional.

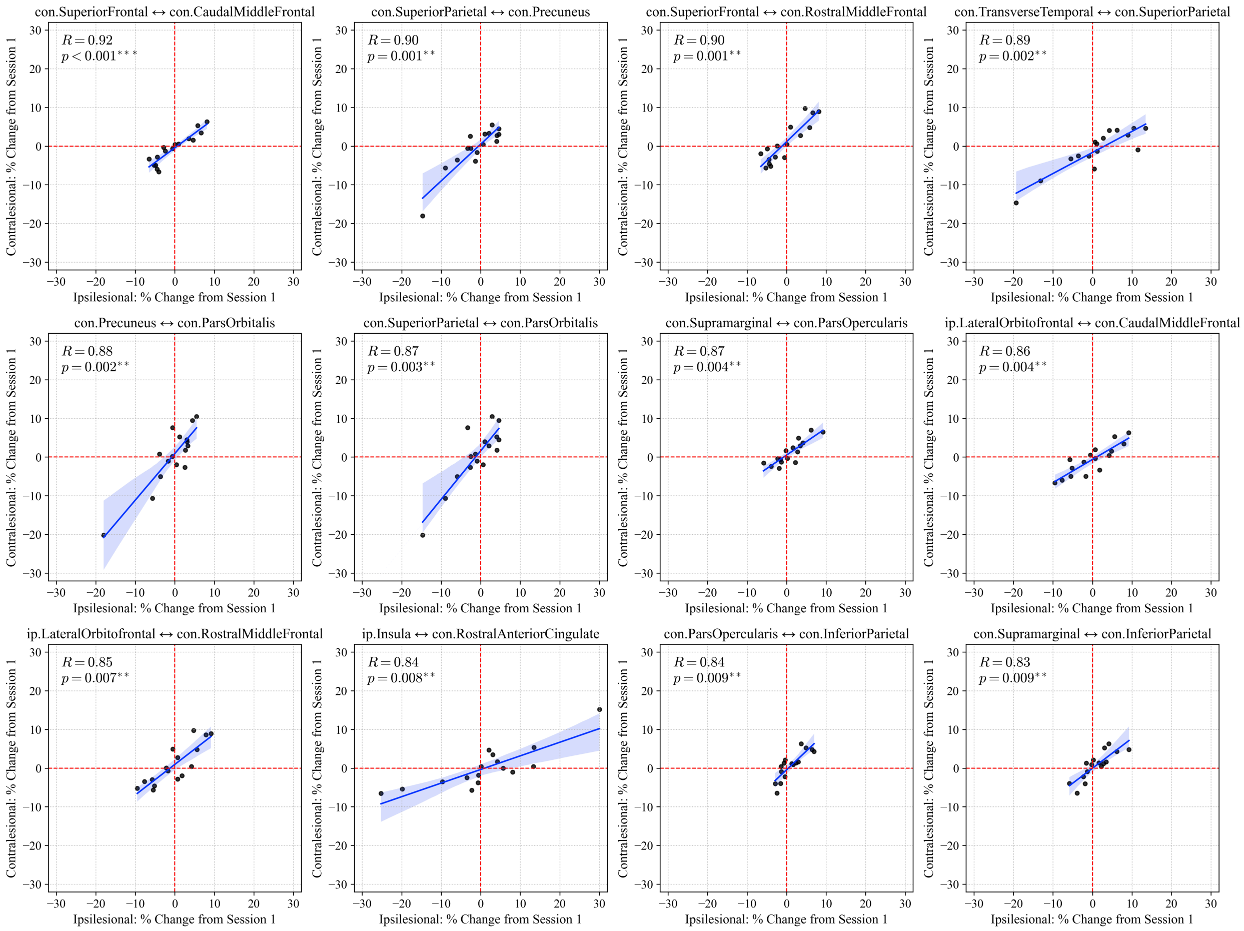


**Figure S12. Non-homologous and homologous region pairs with robust and significant correlations for cortical thickness at 12 months from baseline.** The top 12 significant region pairs (i.e., edges) are displayed. Blue shading indicates Session 3, 12 months from baseline. Scatter plots with the regression line and 95% confidence interval are shown. The p-values displayed are following FDR correction using the BH procedure. * $p < 0.05$, ** $p < 0.01$, *** $p < 0.001$. CON, contralesional; IP, ipsilesional.

**Table S1.**
**Detailed inclusion and exclusion criteria.**

**Study Inclusion Criteria**

- Age > 3 years and < 19 years at brain vascular malformation diagnosis or hemorrhagic stroke
- Screening/enrollment date is occurring prior to the 6 month follow-up visit after hemorrhagic stroke onset or treatment of brain vascular malformation with surgical resection or gamma knife
- Parent/guardian consent & patient assent obtainable (when appropriate)

**Study Exclusion Criteria**

- Patient's brain vascular malformation will be treated with embolization only
- Hemorrhage deemed to be caused by trauma
- Hemorrhagic transformation (intraparenchymal hemorrhage after ischemic infarction)
- Hemorrhage due to cerebral venous thrombosis
- Isolated subdural, epidural hemorrhage, or subarachnoid hemorrhage
- Hemorrhage deemed secondary to surgery or mechanical device (e.g., ECMO)
- Isolated brainstem hemorrhagic stroke/lesion
- Contraindication for MRI (e.g., non-MRI compatible implants, metallic foreign body, or other clinical concern for safety related to MRI or large magnets)
- Pregnancy
- Patient deemed by investigators or clinicians to be unable to undergo additional research sequences or follow-up visits due to safety concerns, quality of data, or other situations
- Lack of parent/guardian available for consent

*Note.* ECMO, extracorporeal membrane oxygenation.

**Table S2.**
**Detailed Patient Demographics, Lesion Characteristics, and Treatment Type.**

| Patient (n=20) | Sex | Side of Lesion | Location of Lesion | Diagnosis | Treatment Type | Number of MRI Sessions |
|---|---|---|---|---|---|---|
| 1 | F | Left | Occipital (Lingual) | AVM | Gamma Knife | *2* |
| 2 | M | Left | Perirolandic | AVM | AVM Resection | 3 |
| 3 | M | Left | Cerebellar | AVM | AVM Resection | 3 |
| 4 | M | Left | Temporal (Anterior) | AVM | AVM Resection and Embolization | 3 |
| 5 | F | *Right* | Paracentral Lobule | AVM | Gamma Knife | *2* |
| 6 | F | Left | Cerebellar (Vermian) | AVM | AVM Resection and Gamma Knife | 3 |
| 7 | M | Left | Temporal (Anterior) | AVM | AVM Resection | 3 |
| 8 | F | Left | Perirolandic/Insular/Temporal Opercular | AVM | AVM Resection | 3 |
| 9 | F | Left | Cerebellar | AVM | AVM Resection | 3 |
| 10 | M | Left | Perirolandic | AVM | AVM Resection | 3 |
| 11 | F | Left | Temporal (Anterior Superior Temporal Gyrus) | AVM | AVM Resection and Embolization | 3 |
| 12 | M | Left | Parietal | AVM | AVM Resection | 3 |
| 13 | M | Left | Temporal (Anterior to Mid Superior Temporal Gyrus) | AVM | AVM Resection | 3 |
| 14 | M | Left | Perirolandic/Insular | *Aneurysm* | Aneurysm Embolization and Clipping | 3 |
| 15 | F | *Right* | Temporal Opercular/Insular | AVM | AVM Resection | 3 |
| 16 | F | Left | Temporal (Medial) | AVM | Gamma Knife | 3 |
| 17 | F | Left | Temporoparietal | AVM | AVM Resection | 3 |
| 18 | F | Left | Inferior Temporal Gyrus | *Unknown VM* | *N/A* | *2* |
| 19 | F | Left | Temporoparietal Junction (TPJ) | *CM* | CM Resection | *2* |
| 20 | M | Left | Frontoparietal | AVM | AVM Resection | 3 |

*Note.* AVM, arteriovenous malformation; CM, cavernous malformation/cavernoma.

**Table S3A.**
**Volume changes in brain tissue segmentation over 3 sessions 6 months apart following pediatric ICH.**

| Hemisphere | Region | Session Comparison | Mean1 (SD1) | Mean2 (SD2) | Mann-Whitney U | p-value | Cohen's *d* |
|---|---|---|---|---|---|---|---|
| Both | CerebralCortex | 1 vs 2 | 0.00 (0.00) | -0.04 (4.05) | 160.00 | 2.534E-01 | 0.01 |
| | | 1 vs 3 | 0.00 (0.00) | -1.35 (3.76) | 200.00 | 1.672E-01 | 0.54 |
| | | 2 vs 3 | -0.04 (4.05) | -1.35 (3.76) | 203.00 | 1.76E-01 | 0.33 |
| | CerebralWM | 1 vs 2 | 0.00 (0.00) | -2.00 (3.10) | 300.00 | **4.016E-03**** | **0.91** |
| | | 1 vs 3 | 0.00 (0.00) | -2.70 (4.01) | 220.00 | **3.747E-02*** | **1.01** |
| | | 2 vs 3 | -2.00 (3.10) | -2.70 (4.01) | 170.00 | 7.623E-01 | 0.20 |
| | SubCortGray | 1 vs 2 | 0.00 (0.00) | -2.15 (3.93) | 300.00 | **4.016E-03**** | **0.77** |
| | | 1 vs 3 | 0.00 (0.00) | -4.15 (4.22) | 300.00 | **1.071E-06***** | **1.48** |
| | | 2 vs 3 | -2.15 (3.93) | -4.15 (4.22) | 217.00 | 7.206E-02 | 0.49 |
| | TotalGray | 1 vs 2 | 0.00 (0.00) | -0.28 (3.46) | 180.00 | 5.729E-01 | 0.12 |
| | | 1 vs 3 | 0.00 (0.00) | -1.59 (3.05) | 240.00 | **5.435E-03**** | **0.79** |
| | | 2 vs 3 | -0.28 (3.46) | -1.59 (3.05) | 205.00 | 1.566E-01 | 0.40 |
| Contralesional | CerebralCortex | 1 vs 2 | 0.00 (0.00) | 1.58 (4.68) | 100.00 | **4.016E-03**** | **-0.48** |
| | | 1 vs 3 | 0.00 (0.00) | 0.78 (4.21) | 140.00 | 4.953E-01 | -0.28 |
| | | 2 vs 3 | 1.58 (4.68) | 0.78 (4.21) | 178.00 | 5.774E-01 | 0.18 |
| | CerebralWM | 1 vs 2 | 0.00 (0.00) | -0.22 (2.15) | 180.00 | 5.729E-01 | 0.14 |
| | | 1 vs 3 | 0.00 (0.00) | 0.23 (1.85) | 140.00 | 4.953E-01 | -0.19 |
| | | 2 vs 3 | -0.22 (2.15) | 0.23 (1.85) | 145.00 | 6.444E-01 | -0.22 |
| | SubCortGray | 1 vs 2 | 0.00 (0.00) | 0.38 (2.20) | 180.00 | 5.729E-01 | -0.25 |
| | | 1 vs 3 | 0.00 (0.00) | -0.46 (2.17) | 180.00 | 4.953E-01 | 0.32 |
| | | 2 vs 3 | 0.38 (2.20) | -0.46 (2.17) | 192.00 | 3.159E-01 | 0.39 |
| | TotalGray | 1 vs 2 | 0.00 (0.00) | 1.39 (4.08) | 120.00 | **2.152E-02*** | **-0.48** |
| | | 1 vs 3 | 0.00 (0.00) | 0.48 (3.66) | 140.00 | 4.953E-01 | -0.20 |
| | | 2 vs 3 | 1.39 (4.08) | 0.48 (3.66) | 182.00 | 4.937E-01 | 0.23 |
| Ipsilesional | CerebralCortex | 1 vs 2 | 0.00 (0.00) | -1.65 (4.75) | 240.00 | 2.534E-01 | 0.49 |
| | | 1 vs 3 | 0.00 (0.00) | -3.48 (4.86) | 220.00 | **3.747E-02*** | **1.08** |
| | | 2 vs 3 | -1.65 (4.75) | -3.48 (4.86) | 197.00 | 2.452E-01 | 0.38 |
| | CerebralWM | 1 vs 2 | 0.00 (0.00) | -3.80 (4.78) | 300.00 | **4.016E-03**** | **1.12** |
| | | 1 vs 3 | 0.00 (0.00) | -5.70 (7.01) | 220.00 | **3.747E-02*** | **1.22** |
| | | 2 vs 3 | -3.80 (4.78) | -5.70 (7.01) | 176.00 | 6.217E-01 | 0.32 |
| | SubCortGray | 1 vs 2 | 0.00 (0.00) | -4.92 (7.85) | 320.00 | **5.499E-04***** | **0.89** |
| | | 1 vs 3 | 0.00 (0.00) | -8.00 (8.90) | 300.00 | **1.071E-06***** | **1.35** |
| | | 2 vs 3 | -4.92 (7.85) | -8.00 (8.90) | 207.00 | 1.388E-01 | 0.37 |
| | TotalGray | 1 vs 2 | 0.00 (0.00) | -1.94 (3.96) | 260.00 | 8.536E-02 | 0.69 |
| | | 1 vs 3 | 0.00 (0.00) | -3.70 (3.78) | 280.00 | **2.931E-05***** | **1.47** |
| | | 2 vs 3 | -1.94 (3.96) | -3.70 (3.78) | 206.00 | 1.475E-01 | 0.45 |

*Note.* Summary statistics—including the mean and standard deviation for each group, p-value following Mann-Whitney U tests, and effect size indicated by Cohen's *d*—are reported. Statistical significance is indicated by asterisks: * $p < 0.05$, ** $p < 0.01$, and *** $p < 0.001$. ICH, intracerebral hemorrhage; WM, white matter.

**Table S3B.**

***Volumes normalized by eTIV:* Volume changes in brain tissue segmentation over 3 sessions 6 months apart following pediatric ICH.**

| Hemisphere | Region | Session Comparison | Mean1 (SD1) | Mean2 (SD2) | Mann-Whitney U | p-value | Cohen's *d* |
|---|---|---|---|---|---|---|---|
| Both | CerebralCortex | 1 vs 2 | 0.00 (0.00) | 1.23 (3.10) | 75.00 | 1.01E-01 | -0.56 |
| | | 1 vs 3 | 0.00 (0.00) | 0.31 (3.83) | 90.00 | 1E+00 | -0.12 |
| | | 2 vs 3 | 1.23 (3.10) | 0.31 (3.83) | 108.00 | 3.932E-01 | 0.27 |
| | CerebralWM | 1 vs 2 | 0.00 (0.00) | -0.97 (3.26) | 120.00 | 7.563E-01 | 0.42 |
| | | 1 vs 3 | 0.00 (0.00) | -0.90 (4.05) | 90.00 | 1E+00 | 0.33 |
| | | 2 vs 3 | -0.97 (3.26) | -0.90 (4.05) | 84.00 | 7.884E-01 | -0.02 |
| | SubCortGray | 1 vs 2 | 0.00 (0.00) | -1.58 (2.76) | 165.00 | **2.115E-02*** | **0.81** |
| | | 1 vs 3 | 0.00 (0.00) | -2.35 (3.80) | 120.00 | 1.139E-01 | 0.93 |
| | | 2 vs 3 | -1.58 (2.76) | -2.35 (3.80) | 101.00 | 6.084E-01 | 0.23 |
| | TotalGray | 1 vs 2 | 0.00 (0.00) | 1.13 (2.96) | 75.00 | 1.01E-01 | -0.54 |
| | | 1 vs 3 | 0.00 (0.00) | 0.46 (3.24) | 90.00 | 1E+00 | -0.21 |
| | | 2 vs 3 | 1.13 (2.96) | 0.46 (3.24) | 108.00 | 3.932E-01 | 0.22 |
| Contralesional | CerebralCortex | 1 vs 2 | 0.00 (0.00) | 2.99 (4.33) | 45.00 | **2.976E-03**** | **-0.98** |
| | | 1 vs 3 | 0.00 (0.00) | 2.55 (4.80) | 60.00 | 1.139E-01 | -0.80 |
| | | 2 vs 3 | 2.99 (4.33) | 2.55 (4.80) | 98.00 | 7.144E-01 | 0.10 |
| | CerebralWM | 1 vs 2 | 0.00 (0.00) | 0.98 (2.55) | 90.00 | 3.294E-01 | -0.54 |
| | | 1 vs 3 | 0.00 (0.00) | 2.19 (2.44) | 45.00 | **1.709E-02*** | **-1.35** |
| | | 2 vs 3 | 0.98 (2.55) | 2.19 (2.44) | 66.00 | 2.515E-01 | -0.48 |
| | SubCortGray | 1 vs 2 | 0.00 (0.00) | 1.05 (2.34) | 75.00 | 1.01E-01 | -0.64 |
| | | 1 vs 3 | 0.00 (0.00) | 1.37 (2.26) | 75.00 | 4.371E-01 | -0.91 |
| | | 2 vs 3 | 1.05 (2.34) | 1.37 (2.26) | 84.00 | 7.884E-01 | -0.13 |
| | TotalGray | 1 vs 2 | 0.00 (0.00) | 2.79 (4.10) | 45.00 | **2.976E-03**** | **-0.96** |
| | | 1 vs 3 | 0.00 (0.00) | 2.52 (4.18) | 60.00 | 1.139E-01 | -0.91 |
| | | 2 vs 3 | 2.79 (4.10) | 2.52 (4.18) | 94.00 | 8.644E-01 | 0.07 |
| Ipsilesional | CerebralCortex | 1 vs 2 | 0.00 (0.00) | -0.52 (3.31) | 105.00 | 7.563E-01 | 0.22 |
| | | 1 vs 3 | 0.00 (0.00) | -1.92 (4.25) | 120.00 | 1.139E-01 | 0.68 |
| | | 2 vs 3 | -0.52 (3.31) | -1.92 (4.25) | 102.00 | 5.747E-01 | 0.37 |
| | CerebralWM | 1 vs 2 | 0.00 (0.00) | -2.92 (4.61) | 150.00 | 1.01E-01 | 0.90 |
| | | 1 vs 3 | 0.00 (0.00) | -4.02 (6.66) | 120.00 | 1.139E-01 | 0.91 |
| | | 2 vs 3 | -2.92 (4.61) | -4.02 (6.66) | 93.00 | 9.029E-01 | 0.20 |
| | SubCortGray | 1 vs 2 | 0.00 (0.00) | -4.66 (5.91) | 225.00 | **6.866E-07***** | **1.12** |
| | | 1 vs 3 | 0.00 (0.00) | -6.47 (7.41) | 165.00 | **6.54E-05***** | **1.32** |
| | | 2 vs 3 | -4.66 (5.91) | -6.47 (7.41) | 115.00 | 2.319E-01 | 0.27 |
| | TotalGray | 1 vs 2 | 0.00 (0.00) | -0.57 (2.95) | 120.00 | 7.563E-01 | 0.27 |
| | | 1 vs 3 | 0.00 (0.00) | -1.68 (3.45) | 120.00 | 1.139E-01 | 0.73 |
| | | 2 vs 3 | -0.57 (2.95) | -1.68 (3.45) | 108.00 | 3.932E-01 | 0.35 |

*Note.* Summary statistics—including the mean and standard deviation for each group, p-value following Mann-Whitney U tests, and effect size indicated by Cohen's *d*—are reported. Session 2 ($n$ = 15) and Session 3 ($n$ = 12) due to the exclusion of 5 participants with poor eTIV measurements. Statistical significance is indicated by asterisks: * $p < 0.05$, ** $p < 0.01$, and *** $p < 0.001$. eTIV, estimated total intracranial volume; ICH, intracerebral hemorrhage; WM, white matter.

**Table S4.**

**Cortical thickness changes of brain hemispheres over 3 sessions 6 months apart following pediatric ICH.**

| Hemisphere | Session Comparison | Mean1 (SD1) | Mean2 (SD2) | Mann-Whitney U | p-value | Cohen's *d* |
|---|---|---|---|---|---|---|
| **Both** | 1 vs 2 | 0.00 (0.00) | 0.34 (2.65) | 140.00 | 8.536E-02 | -0.18 |
| | 1 vs 3 | 0.00 (0.00) | -0.47 (2.76) | 180.00 | 4.953E-01 | 0.26 |
| | 2 vs 3 | 0.34 (2.65) | -0.47 (2.76) | 198.00 | 2.325E-01 | 0.30 |
| **Contralesional** | 1 vs 2 | 0.00 (0.00) | 0.73 (3.18) | 100.00 | **4.016E-03**** | **-0.33** |
| | 1 vs 3 | 0.00 (0.00) | 0.26 (3.16) | 140.00 | 4.953E-01 | -0.12 |
| | 2 vs 3 | 0.73 (3.18) | 0.26 (3.16) | 187.00 | 3.989E-01 | 0.15 |
| **Ipsilesional** | 1 vs 2 | 0.00 (0.00) | -0.05 (2.83) | 220.00 | 5.729E-01 | 0.03 |
| | 1 vs 3 | 0.00 (0.00) | -1.20 (3.10) | 200.00 | 1.672E-01 | 0.58 |
| | 2 vs 3 | -0.05 (2.83) | -1.20 (3.10) | 195.00 | 2.721E-01 | 0.39 |

*Note.* Summary statistics—including the mean and standard deviation for each group, p-value following Mann-Whitney U tests, and effect size indicated by Cohen's *d*—are reported. Statistical significance is indicated by asterisks: * $p < 0.05$, ** $p < 0.01$, and *** $p < 0.001$. ICH, intracerebral hemorrhage.

**Table S5A.**
**Volume changes in brain lobes over 3 sessions 6 months apart following pediatric ICH.**

| Hemisphere | Region | Session Comparison | Mean1 (SD1) | Mean2 (SD2) | Mann-Whitney U | p-value | Cohen's *d* |
|---|---|---|---|---|---|---|---|
| **Both** | Frontal | 1 vs 2 | 0.00 (0.00) | -0.18 (6.38) | 140.00 | 8.536E-02 | 0.04 |
| | | 1 vs 3 | 0.00 (0.00) | -1.06 (6.90) | 160.00 | 1E+00 | 0.23 |
| | | 2 vs 3 | -0.18 (6.38) | -1.06 (6.90) | 168.00 | 8.113E-01 | 0.13 |
| | Parietal | 1 vs 2 | 0.00 (0.00) | -0.46 (6.49) | 200.00 | 1E+00 | 0.10 |
| | | 1 vs 3 | 0.00 (0.00) | -3.28 (4.42) | 240.00 | **5.435E-03**** | **1.12** |
| | | 2 vs 3 | -0.46 (6.49) | -3.28 (4.42) | 213.00 | 9.465E-02 | 0.50 |
| | LateralTemporal | 1 vs 2 | 0.00 (0.00) | -0.98 (7.26) | 180.00 | 5.729E-01 | 0.19 |
| | | 1 vs 3 | 0.00 (0.00) | -1.50 (7.33) | 160.00 | 1E+00 | 0.31 |
| | | 2 vs 3 | -0.98 (7.26) | -1.50 (7.33) | 169.00 | 7.867E-01 | 0.07 |
| | MedialTemporal | 1 vs 2 | 0.00 (0.00) | 3.14 (8.08) | 160.00 | 2.534E-01 | -0.55 |
| | | 1 vs 3 | 0.00 (0.00) | 1.75 (7.39) | 120.00 | 1.672E-01 | -0.36 |
| | | 2 vs 3 | 3.14 (8.08) | 1.75 (7.39) | 169.00 | 7.867E-01 | 0.18 |
| | Occipital | 1 vs 2 | 0.00 (0.00) | 2.05 (5.07) | 120.00 | **2.152E-02*** | **-0.57** |
| | | 1 vs 3 | 0.00 (0.00) | 0.81 (5.44) | 180.00 | 4.953E-01 | -0.22 |
| | | 2 vs 3 | 2.05 (5.07) | 0.81 (5.44) | 194.00 | 2.862E-01 | 0.24 |
| **Contra-lesional** | Frontal | 1 vs 2 | 0.00 (0.00) | 0.82 (7.99) | 120.00 | **2.152E-02*** | **-0.15** |
| | | 1 vs 3 | 0.00 (0.00) | 0.78 (6.50) | 160.00 | 1E+00 | -0.18 |
| | | 2 vs 3 | 0.82 (7.99) | 0.78 (6.50) | 175.00 | 6.444E-01 | 0.01 |
| | Parietal | 1 vs 2 | 0.00 (0.00) | 3.05 (8.73) | 60.00 | **5.496E-05***** | **-0.49** |
| | | 1 vs 3 | 0.00 (0.00) | 0.28 (6.45) | 140.00 | 4.953E-01 | -0.06 |
| | | 2 vs 3 | 3.05 (8.73) | 0.28 (6.45) | 203.00 | 1.76E-01 | 0.35 |
| | LateralTemporal | 1 vs 2 | 0.00 (0.00) | 1.50 (5.32) | 140.00 | 8.536E-02 | -0.40 |
| | | 1 vs 3 | 0.00 (0.00) | 1.29 (4.29) | 100.00 | **3.747E-02*** | **-0.45** |
| | | 2 vs 3 | 1.50 (5.32) | 1.29 (4.29) | 160.00 | 1E+00 | 0.04 |
| | MedialTemporal | 1 vs 2 | 0.00 (0.00) | 3.05 (5.81) | 120.00 | **2.152E-02*** | **-0.74** |
| | | 1 vs 3 | 0.00 (0.00) | 2.71 (5.38) | 100.00 | **3.747E-02*** | **-0.76** |
| | | 2 vs 3 | 3.05 (5.81) | 2.71 (5.38) | 160.00 | 1E+00 | 0.06 |
| | Occipital | 1 vs 2 | 0.00 (0.00) | 2.44 (4.39) | 100.00 | **4.016E-03**** | **-0.79** |
| | | 1 vs 3 | 0.00 (0.00) | 0.61 (3.30) | 160.00 | 1E+00 | -0.28 |
| | | 2 vs 3 | 2.44 (4.39) | 0.61 (3.30) | 196.00 | 2.584E-01 | 0.46 |
| **Ipsilesional** | Frontal | 1 vs 2 | 0.00 (0.00) | -1.07 (6.71) | 200.00 | 1E+00 | 0.23 |
| | | 1 vs 3 | 0.00 (0.00) | -2.83 (9.22) | 200.00 | 1.672E-01 | 0.46 |
| | | 2 vs 3 | -1.07 (6.71) | -2.83 (9.22) | 177.00 | 5.994E-01 | 0.22 |
| | Parietal | 1 vs 2 | 0.00 (0.00) | -3.81 (8.98) | 240.00 | 2.534E-01 | 0.60 |
| | | 1 vs 3 | 0.00 (0.00) | -6.78 (8.38) | 260.00 | **5.025E-04***** | **1.22** |
| | | 2 vs 3 | -3.81 (8.98) | -6.78 (8.38) | 202.00 | 1.864E-01 | 0.34 |
| | LateralTemporal | 1 vs 2 | 0.00 (0.00) | -3.39 (12.56) | 240.00 | 2.534E-01 | 0.38 |
| | | 1 vs 3 | 0.00 (0.00) | -4.24 (13.85) | 180.00 | 4.953E-01 | 0.46 |
| | | 2 vs 3 | -3.39 (12.56) | -4.24 (13.85) | 162.00 | 9.619E-01 | 0.06 |
| | MedialTemporal | 1 vs 2 | 0.00 (0.00) | 3.73 (13.20) | 160.00 | 2.534E-01 | -0.40 |
| | | 1 vs 3 | 0.00 (0.00) | 1.15 (11.58) | 160.00 | 1E+00 | -0.15 |
| | | 2 vs 3 | 3.73 (13.20) | 1.15 (11.58) | 176.00 | 6.217E-01 | 0.21 |
| | Occipital | 1 vs 2 | 0.00 (0.00) | 2.08 (10.24) | 180.00 | 5.729E-01 | -0.29 |
| | | 1 vs 3 | 0.00 (0.00) | 1.67 (13.02) | 200.00 | 1.672E-01 | -0.19 |
| | | 2 vs 3 | 2.08 (10.24) | 1.67 (13.02) | 181.00 | 5.14E-01 | 0.04 |

*Note.* Summary statistics—including the mean and standard deviation for each group, p-value following Mann-Whitney U tests, and effect size indicated by Cohen's *d*—are reported. Statistical significance is indicated by asterisks: * $p < 0.05$, ** $p < 0.01$, and *** $p < 0.001$. ICH, intracerebral hemorrhage.

**Table S5B.**

***Volumes normalized by eTIV:*** **Volume changes in brain lobes over 3 sessions 6 months apart following pediatric ICH.**

| Hemisphere | Region | Session Comparison | Mean1 (SD1) | Mean2 (SD2) | Mann-Whitney U | p-value | Cohen's *d* |
|---|---|---|---|---|---|---|---|
| **Both** | Frontal | 1 vs 2 | 0.00 (0.00) | 1.83 (4.63) | 52.50 | **7.387E-03**** | **-0.56** |
| | | 1 vs 3 | 0.00 (0.00) | 0.95 (7.42) | 75.00 | 4.371E-01 | -0.19 |
| | | 2 vs 3 | 1.83 (4.63) | 0.95 (7.42) | 89.00 | 9.805E-01 | 0.15 |
| | Parietal | 1 vs 2 | 0.00 (0.00) | 1.07 (4.90) | 67.50 | **4.512E-02*** | **-0.31** |
| | | 1 vs 3 | 0.00 (0.00) | -1.38 (4.56) | 97.50 | 7.012E-01 | 0.46 |
| | | 2 vs 3 | 1.07 (4.90) | -1.38 (4.56) | 113.50 | 2.615E-01 | 0.51 |
| | LateralTemporal | 1 vs 2 | 0.00 (0.00) | -0.72 (8.08) | 105.00 | 7.483E-01 | 0.13 |
| | | 1 vs 3 | 0.00 (0.00) | -0.74 (8.04) | 90.00 | 1E+00 | 0.14 |
| | | 2 vs 3 | -0.72 (8.08) | -0.74 (8.04) | 94.00 | 8.641E-01 | 0.00 |
| | MedialTemporal | 1 vs 2 | 0.00 (0.00) | 3.54 (8.05) | 90.00 | 2.604E-01 | -0.62 |
| | | 1 vs 3 | 0.00 (0.00) | 1.68 (8.59) | 75.00 | 3.809E-01 | -0.29 |
| | | 2 vs 3 | 3.54 (8.05) | 1.68 (8.59) | 96.50 | 7.637E-01 | 0.23 |
| | Occipital | 1 vs 2 | 0.00 (0.00) | 2.22 (6.35) | 90.00 | 3.131E-01 | -0.49 |
| | | 1 vs 3 | 0.00 (0.00) | 4.01 (6.61) | 45.00 | **2.856E-03**** | **-0.92** |
| | | 2 vs 3 | 2.22 (6.35) | 4.01 (6.61) | 74.00 | 4.434E-01 | -0.28 |
| **Contra-lesional** | Frontal | 1 vs 2 | 0.00 (0.00) | 3.13 (6.94) | 37.50 | **7.946E-04***** | **-0.64** |
| | | 1 vs 3 | 0.00 (0.00) | 3.60 (6.56) | 67.50 | 2.278E-01 | -0.83 |
| | | 2 vs 3 | 3.13 (6.94) | 3.60 (6.56) | 92.50 | 9.222E-01 | -0.07 |
| | Parietal | 1 vs 2 | 0.00 (0.00) | 4.67 (9.04) | 30.00 | **2.778E-04***** | **-0.73** |
| | | 1 vs 3 | 0.00 (0.00) | 1.59 (7.36) | 52.50 | **3.141E-02*** | **-0.32** |
| | | 2 vs 3 | 4.67 (9.04) | 1.59 (7.36) | 112.00 | 2.934E-01 | 0.37 |
| | LateralTemporal | 1 vs 2 | 0.00 (0.00) | 1.61 (5.79) | 105.00 | 7.368E-01 | -0.39 |
| | | 1 vs 3 | 0.00 (0.00) | 2.58 (5.99) | 67.50 | 1.637E-01 | -0.65 |
| | | 2 vs 3 | 1.61 (5.79) | 2.58 (5.99) | 81.50 | 6.907E-01 | -0.16 |
| | MedialTemporal | 1 vs 2 | 0.00 (0.00) | 2.65 (8.39) | 90.00 | 2.182E-01 | -0.45 |
| | | 1 vs 3 | 0.00 (0.00) | 2.53 (7.93) | 75.00 | 2.517E-01 | -0.48 |
| | | 2 vs 3 | 2.65 (8.39) | 2.53 (7.93) | 89.50 | 1E+00 | 0.01 |
| | Occipital | 1 vs 2 | 0.00 (0.00) | 2.87 (5.41) | 75.00 | 5.83E-02 | -0.75 |
| | | 1 vs 3 | 0.00 (0.00) | 4.00 (4.74) | 45.00 | **2.848E-03**** | **-1.27** |
| | | 2 vs 3 | 2.87 (5.41) | 4.00 (4.74) | 75.50 | 4.741E-01 | -0.22 |
| **Ipsilesional** | Frontal | 1 vs 2 | 0.00 (0.00) | 0.69 (5.60) | 90.00 | 3.294E-01 | -0.17 |
| | | 1 vs 3 | 0.00 (0.00) | -1.57 (10.31) | 97.50 | 7.011E-01 | 0.23 |
| | | 2 vs 3 | 0.69 (5.60) | -1.57 (10.31) | 93.00 | 9.029E-01 | 0.28 |
| | Parietal | 1 vs 2 | 0.00 (0.00) | -2.34 (6.09) | 135.00 | 3.294E-01 | 0.54 |
| | | 1 vs 3 | 0.00 (0.00) | -4.32 (6.46) | 135.00 | **1.709E-02*** | **1.01** |
| | | 2 vs 3 | -2.34 (6.09) | -4.32 (6.46) | 110.50 | 3.283E-01 | 0.32 |
| | LateralTemporal | 1 vs 2 | 0.00 (0.00) | -2.88 (14.46) | 112.50 | 1E+00 | 0.28 |
| | | 1 vs 3 | 0.00 (0.00) | -3.91 (15.79) | 90.00 | 1E+00 | 0.37 |
| | | 2 vs 3 | -2.88 (14.46) | -3.91 (15.79) | 92.00 | 9.415E-01 | 0.07 |
| | MedialTemporal | 1 vs 2 | 0.00 (0.00) | 5.30 (13.26) | 82.50 | 1.157E-01 | -0.57 |
| | | 1 vs 3 | 0.00 (0.00) | 1.33 (11.88) | 75.00 | 3.309E-01 | -0.17 |
| | | 2 vs 3 | 5.30 (13.26) | 1.33 (11.88) | 98.50 | 6.777E-01 | 0.31 |
| | Occipital | 1 vs 2 | 0.00 (0.00) | 2.38 (14.08) | 127.50 | 4.735E-01 | -0.24 |
| | | 1 vs 3 | 0.00 (0.00) | 5.19 (17.06) | 82.50 | 5.315E-01 | -0.46 |
| | | 2 vs 3 | 2.38 (14.08) | 5.19 (17.06) | 70.50 | 3.175E-01 | -0.18 |

*Note.* Summary statistics—including the mean and standard deviation for each group, p-value following Mann-Whitney U tests, and effect size indicated by Cohen's *d*—are reported. Session 2 ($n = 15$) and Session 3 ($n = 12$) due to the exclusion of 5 participants with poor eTIV measurements. Statistical significance is indicated by asterisks: * $p < 0.05$, ** $p < 0.01$, and *** $p < 0.001$. eTIV, estimated total intracranial volume; ICH, intracerebral hemorrhage.

**Table S6.**
**Cortical thickness changes of brain lobes over 3 sessions 6 months apart following pediatric ICH.**

| Hemisphere | Region | Session Comparison | Mean1 (SD1) | Mean2 (SD2) | Mann-Whitney U | p-value | Cohen's *d* |
|---|---|---|---|---|---|---|---|
| **Both** | Frontal | 1 vs 2 | 0.00 (0.00) | 0.65 (3.84) | 160.00 | 2.534E-01 | -0.24 |
| | | 1 vs 3 | 0.00 (0.00) | -0.27 (3.78) | 200.00 | 1.672E-01 | 0.11 |
| | | 2 vs 3 | 0.65 (3.84) | -0.27 (3.78) | 201.00 | 1.973E-01 | 0.24 |
| | Parietal | 1 vs 2 | 0.00 (0.00) | 0.06 (3.45) | 190.00 | 7.811E-01 | -0.02 |
| | | 1 vs 3 | 0.00 (0.00) | -1.77 (4.07) | 220.00 | **3.747E-02*** | **0.66** |
| | | 2 vs 3 | 0.06 (3.45) | -1.77 (4.07) | 199.00 | 2.203E-01 | 0.49 |
| | LateralTemporal | 1 vs 2 | 0.00 (0.00) | 1.16 (3.81) | 160.00 | 2.534E-01 | -0.43 |
| | | 1 vs 3 | 0.00 (0.00) | 0.70 (3.54) | 160.00 | 1E+00 | -0.30 |
| | | 2 vs 3 | 1.16 (3.81) | 0.70 (3.54) | 176.00 | 6.217E-01 | 0.13 |
| | MedialTemporal | 1 vs 2 | 0.00 (0.00) | 0.79 (3.78) | 200.00 | 1E+00 | -0.30 |
| | | 1 vs 3 | 0.00 (0.00) | 1.80 (4.10) | 120.00 | 1.672E-01 | -0.66 |
| | | 2 vs 3 | 0.79 (3.78) | 1.80 (4.10) | 137.00 | 4.738E-01 | -0.26 |
| | Occipital | 1 vs 2 | 0.00 (0.00) | -0.37 (3.89) | 240.00 | 2.534E-01 | 0.13 |
| | | 1 vs 3 | 0.00 (0.00) | -0.87 (3.17) | 180.00 | 4.953E-01 | 0.41 |
| | | 2 vs 3 | -0.37 (3.89) | -0.87 (3.17) | 174.00 | 6.674E-01 | 0.14 |
| **Contra-lesional** | Frontal | 1 vs 2 | 0.00 (0.00) | 1.16 (4.24) | 100.00 | **4.016E-03**** | **-0.39** |
| | | 1 vs 3 | 0.00 (0.00) | 0.20 (4.34) | 180.00 | 4.953E-01 | -0.07 |
| | | 2 vs 3 | 1.16 (4.24) | 0.20 (4.34) | 197.00 | 2.452E-01 | 0.22 |
| | Parietal | 1 vs 2 | 0.00 (0.00) | 1.11 (3.76) | 120.00 | **2.152E-02*** | **-0.42** |
| | | 1 vs 3 | 0.00 (0.00) | 0.32 (4.10) | 140.00 | 4.953E-01 | -0.12 |
| | | 2 vs 3 | 1.11 (3.76) | 0.32 (4.10) | 176.00 | 6.217E-01 | 0.20 |
| | LateralTemporal | 1 vs 2 | 0.00 (0.00) | 1.13 (4.41) | 140.00 | 8.536E-02 | -0.36 |
| | | 1 vs 3 | 0.00 (0.00) | 0.85 (3.72) | 140.00 | 4.953E-01 | -0.34 |
| | | 2 vs 3 | 1.13 (4.41) | 0.85 (3.72) | 173.50 | 6.79E-01 | 0.07 |
| | MedialTemporal | 1 vs 2 | 0.00 (0.00) | 0.34 (4.60) | 170.00 | 3.883E-01 | -0.10 |
| | | 1 vs 3 | 0.00 (0.00) | 1.73 (4.73) | 120.00 | 1.672E-01 | -0.55 |
| | | 2 vs 3 | 0.34 (4.60) | 1.73 (4.73) | 132.00 | 3.813E-01 | -0.30 |
| | Occipital | 1 vs 2 | 0.00 (0.00) | 0.62 (3.75) | 120.00 | **2.152E-02*** | **-0.23** |
| | | 1 vs 3 | 0.00 (0.00) | 0.28 (3.10) | 120.00 | 1.672E-01 | -0.14 |
| | | 2 vs 3 | 0.62 (3.75) | 0.28 (3.10) | 162.00 | 9.619E-01 | 0.10 |
| **Ipsilesional** | Frontal | 1 vs 2 | 0.00 (0.00) | 0.17 (4.30) | 260.00 | 8.536E-02 | -0.05 |
| | | 1 vs 3 | 0.00 (0.00) | -0.73 (4.02) | 240.00 | **5.435E-03**** | **0.27** |
| | | 2 vs 3 | 0.17 (4.30) | -0.73 (4.02) | 198.00 | 2.325E-01 | 0.21 |
| | Parietal | 1 vs 2 | 0.00 (0.00) | -0.96 (4.88) | 240.00 | 2.534E-01 | 0.28 |
| | | 1 vs 3 | 0.00 (0.00) | -3.84 (6.24) | 240.00 | **5.435E-03**** | **0.93** |
| | | 2 vs 3 | -0.96 (4.88) | -3.84 (6.24) | 201.00 | 1.973E-01 | 0.52 |
| | LateralTemporal | 1 vs 2 | 0.00 (0.00) | 1.30 (5.50) | 180.00 | 5.729E-01 | -0.33 |
| | | 1 vs 3 | 0.00 (0.00) | 0.59 (4.68) | 160.00 | 1E+00 | -0.19 |
| | | 2 vs 3 | 1.30 (5.50) | 0.59 (4.68) | 176.00 | 6.217E-01 | 0.14 |
| | MedialTemporal | 1 vs 2 | 0.00 (0.00) | 1.42 (6.80) | 200.00 | 1E+00 | -0.30 |
| | | 1 vs 3 | 0.00 (0.00) | 2.07 (6.42) | 120.00 | 1.672E-01 | -0.49 |
| | | 2 vs 3 | 1.42 (6.80) | 2.07 (6.42) | 154.00 | 8.61E-01 | -0.10 |
| | Occipital | 1 vs 2 | 0.00 (0.00) | -1.32 (4.76) | 280.00 | **2.152E-02*** | **0.39** |
| | | 1 vs 3 | 0.00 (0.00) | -1.95 (4.51) | 240.00 | **5.435E-03**** | **0.65** |
| | | 2 vs 3 | -1.32 (4.76) | -1.95 (4.51) | 166.00 | 8.61E-01 | 0.14 |

*Note.* Summary statistics—including the mean and standard deviation for each group, p-value following Mann-Whitney U tests, and effect size indicated by Cohen's *d*—are reported. Statistical significance is indicated by asterisks: * $p < 0.05$, ** $p < 0.01$, and *** $p < 0.001$. ICH, intracerebral hemorrhage.

**Table S7A.**
**Volume changes in corpus callosum over 3 sessions 6 months apart following pediatric ICH.**

| Region | Session Comparison | Mean1 (SD1) | Mean2 (SD2) | Mann-Whitney U | p-value | Cohen's *d* |
|---|---|---|---|---|---|---|
| CCAnterior | 1 vs 2 | 0.00 (0.00) | -1.14 (16.24) | 220.00 | 5.729E-01 | 0.10 |
| | 1 vs 3 | 0.00 (0.00) | 3.82 (17.51) | 160.00 | 1E+00 | -0.33 |
| | 2 vs 3 | -1.14 (16.24) | 3.82 (17.51) | 131.00 | 3.642E-01 | -0.29 |
| CCMidAnterior | 1 vs 2 | 0.00 (0.00) | -0.89 (29.01) | 220.00 | 5.729E-01 | 0.04 |
| | 1 vs 3 | 0.00 (0.00) | -14.14 (22.46) | 240.00 | **5.435E-03**** | **0.95** |
| | 2 vs 3 | -0.89 (29.01) | -14.14 (22.46) | 200.00 | 2.086E-01 | 0.50 |
| CCCentral | 1 vs 2 | 0.00 (0.00) | -0.39 (20.47) | 260.00 | 8.536E-02 | 0.03 |
| | 1 vs 3 | 0.00 (0.00) | -6.55 (19.70) | 240.00 | **5.435E-03**** | **0.50** |
| | 2 vs 3 | -0.39 (20.47) | -6.55 (19.70) | 192.00 | 3.159E-01 | 0.31 |
| CCMidPosterior | 1 vs 2 | 0.00 (0.00) | -6.06 (14.74) | 300.00 | **4.016E-03**** | **0.58** |
| | 1 vs 3 | 0.00 (0.00) | -10.07 (13.53) | 260.00 | **5.025E-04***** | **1.12** |
| | 2 vs 3 | -6.06 (14.74) | -10.07 (13.53) | 163.00 | 9.366E-01 | 0.28 |
| CCPosterior | 1 vs 2 | 0.00 (0.00) | -2.64 (8.05) | 260.00 | 8.536E-02 | 0.46 |
| | 1 vs 3 | 0.00 (0.00) | -4.43 (6.48) | 220.00 | **3.747E-02*** | **1.03** |
| | 2 vs 3 | -2.64 (8.05) | -4.43 (6.48) | 177.00 | 5.994E-01 | 0.24 |
| CCTotal | 1 vs 2 | 0.00 (0.00) | -3.10 (7.62) | 280.00 | **2.152E-02*** | **0.57** |
| | 1 vs 3 | 0.00 (0.00) | -6.15 (8.06) | 240.00 | **5.435E-03**** | **1.15** |
| | 2 vs 3 | -3.10 (7.62) | -6.15 (8.06) | 184.00 | 4.544E-01 | 0.39 |

*Note.* Summary statistics—including the mean and standard deviation for each group, p-value following Mann-Whitney U tests, and effect size indicated by Cohen's *d*—are reported. Statistical significance is indicated by asterisks: * $p < 0.05$, ** $p < 0.01$, and *** $p < 0.001$. CC, corpus callosum; ICH, intracerebral hemorrhage.

**Table S7B.**
***Volumes normalized by eTIV:*** **volume changes in corpus callosum over 3 sessions 6 months apart following pediatric ICH.**

| Region | Session Comparison | Mean1 (SD1) | Mean2 (SD2) | Mann-Whitney U | p-value | Cohen's *d* |
|---|---|---|---|---|---|---|
| CCAnterior | 1 vs 2 | 0.00 (0.00) | -0.20 (17.15) | 120.00 | 7.563E-01 | 0.02 |
| | 1 vs 3 | 0.00 (0.00) | 7.31 (20.54) | 90.00 | 1E+00 | -0.54 |
| | 2 vs 3 | -0.20 (17.15) | 7.31 (20.54) | 69.00 | 3.172E-01 | -0.40 |
| CCMidAnterior | 1 vs 2 | 0.00 (0.00) | 3.44 (31.32) | 105.00 | 7.563E-01 | -0.16 |
| | 1 vs 3 | 0.00 (0.00) | -9.14 (19.75) | 135.00 | **1.709E-02*** | **0.70** |
| | 2 vs 3 | 3.44 (31.32) | -9.14 (19.75) | 113.00 | 2.723E-01 | 0.47 |
| CCCentral | 1 vs 2 | 0.00 (0.00) | 3.38 (20.39) | 135.00 | 3.294E-01 | -0.23 |
| | 1 vs 3 | 0.00 (0.00) | 0.11 (20.27) | 90.00 | 1E+00 | -0.01 |
| | 2 vs 3 | 3.38 (20.39) | 0.11 (20.27) | 98.00 | 7.144E-01 | 0.16 |
| CCMidPosterior | 1 vs 2 | 0.00 (0.00) | -6.13 (12.88) | 180.00 | **2.976E-03**** | **0.67** |
| | 1 vs 3 | 0.00 (0.00) | -8.05 (10.98) | 150.00 | **1.43E-03**** | **1.11** |
| | 2 vs 3 | -6.13 (12.88) | -8.05 (10.98) | 85.00 | 8.262E-01 | 0.16 |
| CCPosterior | 1 vs 2 | 0.00 (0.00) | -0.45 (8.37) | 105.00 | 7.563E-01 | 0.08 |
| | 1 vs 3 | 0.00 (0.00) | -1.88 (6.26) | 90.00 | 1E+00 | 0.45 |
| | 2 vs 3 | -0.45 (8.37) | -1.88 (6.26) | 99.00 | 6.783E-01 | 0.19 |
| CCTotal | 1 vs 2 | 0.00 (0.00) | -0.84 (7.40) | 150.00 | 1.01E-01 | 0.16 |
| | 1 vs 3 | 0.00 (0.00) | -2.19 (7.52) | 120.00 | 1.139E-01 | 0.44 |
| | 2 vs 3 | -0.84 (7.40) | -2.19 (7.52) | 86.00 | 8.644E-01 | 0.18 |

*Note.* Summary statistics—including the mean and standard deviation for each group, p-value following Mann-Whitney U tests, and effect size indicated by Cohen's *d*—are reported. Session 2 ($n$ = 15) and Session 3 ($n$ = 12) due to the exclusion of 5 participants with poor eTIV measurements. Statistical significance is indicated by asterisks: * $p < 0.05$, ** $p < 0.01$, and *** $p < 0.001$. eTIV, estimated total intracranial volume; CC, corpus callosum; ICH, intracerebral hemorrhage.

**Table S8A.**

**Volume changes in CSF over 3 sessions 6 months apart following pediatric ICH.**

| Region | Session Comparison | Mean1 (SD1) | Mean2 (SD2) | Mann-Whitney U | p-value | Cohen's *d* |
|---|---|---|---|---|---|---|
| CSF | 1 vs 2 | 0.00 (0.00) | 1.93 (9.49) | 200.00 | 1E+00 | -0.29 |
| | 1 vs 3 | 0.00 (0.00) | 5.78 (8.82) | 100.00 | **3.747E-02*** | **-0.99** |
| | 2 vs 3 | 1.93 (9.49) | 5.78 (8.82) | 120.00 | 2.086E-01 | -0.42 |

*Note.* Summary statistics—including the mean and standard deviation for each group, p-value following Mann-Whitney U tests, and effect size indicated by Cohen's *d*—are reported. Statistical significance is indicated by asterisks: * $p < 0.05$, ** $p < 0.01$, and *** $p < 0.001$. CSF, cerebrospinal fluid; ICH, intracerebral hemorrhage.

**Table S8B.**

***Volumes normalized by eTIV:* volume changes in CSF over 3 sessions 6 months apart following pediatric ICH.**

| Region | Session Comparison | Mean1 (SD1) | Mean2 (SD2) | Mann-Whitney U | p-value | Cohen's *d* |
|---|---|---|---|---|---|---|
| CSF | 1 vs 2 | 0.00 (0.00) | -0.01 (9.38) | 135.00 | 3.294E-01 | 0.00 |
| | 1 vs 3 | 0.00 (0.00) | 6.13 (8.63) | 45.00 | **1.709E-02*** | **-1.07** |
| | 2 vs 3 | -0.01 (9.38) | 6.13 (8.63) | 47.00 | **3.81E-02*** | **-0.68** |

*Note.* Summary statistics—including the mean and standard deviation for each group, p-value following Mann-Whitney U tests, and effect size indicated by Cohen's *d*—are reported. Session 2 ($n = 15$) and Session 3 ($n = 12$) due to the exclusion of 5 participants with poor eTIV measurements. Statistical significance is indicated by asterisks: * $p < 0.05$, ** $p < 0.01$, and *** $p < 0.001$. eTIV, estimated total intracranial volume; CSF, cerebrospinal fluid; ICH, intracerebral hemorrhage.

**Table S9A.**
**Associations between the volume and cortical thickness changes of homologous contralesional and ipsilesional regions and lobes per 3 sessions 6 months apart following pediatric ICH.**

| | | Regions | | | | | Lobes | | | | |
|---|---|---|---|---|---|---|---|---|---|---|---|
| **Metric** | **Session** | ***R*** | ***Slope*** | ***SE*** | ***p*** | ***p (adj)*** | ***R*** | ***Slope*** | ***SE*** | ***p*** | ***p (adj)*** |
| **Volume** | **2 (6m)** | 0.11 | 0.19 | 0.07 | **0.004**** | **0.030*** | 0.26 | 0.42 | 0.14 | **0.004**** | **0.031*** |
| | **3 (12m)** | 0.14 | 0.26 | 0.08 | **0.002**** | **0.017*** | 0.21 | 0.44 | 0.21 | **0.037*** | **0.048*** |
| **Cortical Thickness** | **2 (6m)** | 0.26 | 0.36 | 0.05 | **< 0.001***** | **< 0.001***** | 0.29 | 0.36 | 0.10 | **< 0.001***** | **0.023*** |
| | **3 (12m)** | 0.29 | 0.42 | 0.06 | **< 0.001***** | **< 0.001***** | 0.35 | 0.48 | 0.12 | **< 0.001***** | **< 0.001***** |

*Note.* Pearson correlation coefficients (*R*), slopes, standard errors (*SE*), and significance levels are reported for both regional (*left*) and lobar (*right*) analyses across sessions. Significance was evaluated using non-parametric permutation testing ($n = 5{,}000$). All *p*-values were rounded to three decimal places. * $p < 0.05$, ** $p < 0.01$, and *** $p < 0.001$. ICH, intracerebral hemorrhage.

**Table S9B.**
***Volumes normalized by eTIV:* associations between the volume changes of homologous contralesional and ipsilesional regions and lobes per 3 sessions 6 months apart following pediatric ICH.**

| | | Regions | | | | | Lobes | | | | |
|---|---|---|---|---|---|---|---|---|---|---|---|
| **Metric** | **Session** | ***R*** | ***Slope*** | ***SE*** | ***p*** | ***p (adj)*** | ***R*** | ***Slope*** | ***SE*** | ***p*** | ***p (adj)*** |
| **Volume** | **2 (6m)** | 0.07 | 0.12 | 0.08 | 0.146 | 0.234 | 0.07 | 0.12 | 0.17 | 0.491 | 0.624 |
| | **3 (12m)** | 0.15 | 0.29 | 0.10 | **0.005**** | **0.033*** | 0.13 | 0.25 | 0.22 | 0.259 | 0.164 |

*Note.* Pearson correlation coefficients (*R*), slopes, standard errors (*SE*), and significance levels are reported for both regional (*left*) and lobar (*right*) analyses across sessions. Significance was evaluated using non-parametric permutation testing ($n = 5{,}000$). Session 2 ($n = 15$) and Session 3 ($n = 12$) due to the exclusion of 5 participants with poor eTIV measurements. All *p*-values were rounded to three decimal places. * $p < 0.05$, ** $p < 0.01$, and *** $p < 0.001$. eTIV, estimated total intracranial volume; ICH, intracerebral hemorrhage.

**Table S10.**
**Regions with robust and significant homologous correlations for cortical thickness at 6 and 12 months from baseline.**

| Session | Region | ***R*** | ***p*** | ***p (adj)*** |
|---|---|---|---|---|
| 2 (6 months from Baseline) | SuperiorFrontal | 0.93 | **< 0.001*** ** | **< 0.001*** ** |
| | MedialOrbitofrontal | 0.81 | **< 0.001*** ** | **< 0.001*** ** |
| | Pericalcarine | 0.78 | **< 0.001*** ** | **< 0.001*** ** |
| | RostralMiddleFrontal | 0.77 | **< 0.001*** ** | **< 0.001*** ** |
| | Paracentral | 0.74 | **< 0.001*** ** | **0.001*** ** |
| | LateralOrbitofrontal | 0.71 | **< 0.001*** ** | **0.002** ** |
| | RostralAnteriorCingulate | 0.56 | **0.010** ** | **0.044* ** |
| | LateralOccipital | 0.55 | **0.012* ** | **0.046* ** |
| 3 (12 months from Baseline) | SuperiorFrontal | 0.80 | **< 0.001*** ** | **0.005** ** |
| | LateralOccipital | 0.73 | **0.001*** ** | **0.016* ** |
| | MedialOrbitofrontal | 0.72 | **0.002** ** | **0.016* ** |
| | RostralMiddleFrontal | 0.68 | **0.004** ** | **0.025* ** |
| | Pericalcarine | 0.68 | **0.004** ** | **0.025* ** |
| | PosteriorCingulate | 0.63 | **0.009** ** | **0.039* ** |
| | Insula | 0.63 | **0.009** ** | **0.039* ** |
| | ParsOrbitalis | 0.62 | **0.010** ** | **0.039* ** |
| | LateralOrbitofrontal | 0.60 | **0.014* ** | **0.048* ** |
| | ParsTriangularis | 0.59 | **0.015* ** | **0.048* ** |

*Note.* Only the significant regions of Sessions 2 and 3 are displayed. The adjusted p-values displayed are following FDR correction using the BH procedure. All *p*-values were rounded to three decimal places. * $p < 0.05$, ** $p < 0.01$, *** $p < 0.001$.

**Table S11.**
**Non-homologous and homologous region pairs with robust and significant correlations for cortical thickness at 6 months and 12 months from baseline.**

| Session | Node 1 | Node 2 | *R* | *p* | *p (adj)* |
|---|---|---|---|---|---|
| 2 (6 months from Baseline) | con.SuperiorFrontal | con.CaudalMiddleFrontal | 0.97 | **< 0.001*** ** | **< 0.001*** ** |
| | ip.SuperiorFrontal | con.SuperiorFrontal | 0.93 | **< 0.001*** ** | **< 0.001*** ** |
| | con.Paracentral | con.CaudalMiddleFrontal | 0.91 | **< 0.001*** ** | **< 0.001*** ** |
| | ip.SuperiorFrontal | con.CaudalMiddleFrontal | 0.91 | **< 0.001*** ** | **< 0.001*** ** |
| | con.Supramarginal | con.InferiorParietal | 0.89 | **< 0.001*** ** | **< 0.001*** ** |
| | ip.SuperiorParietal | ip.InferiorParietal | 0.88 | **< 0.001*** ** | **< 0.001*** ** |
| | con.SuperiorFrontal | con.Paracentral | 0.87 | **< 0.001*** ** | **< 0.001*** ** |
| | con.SuperiorTemporal | con.MiddleTemporal | 0.87 | **< 0.001*** ** | **< 0.001*** ** |
| | con.Precuneus | con.CaudalMiddleFrontal | 0.82 | **< 0.001*** ** | **0.002** ** |
| | con.SuperiorFrontal | con.Precuneus | 0.82 | **< 0.001*** ** | **0.002** ** |
| | ip.Pericalcarine | ip.Lingual | 0.82 | **< 0.001*** ** | **0.002** ** |
| | con.RostralMiddleFrontal | con.Precuneus | 0.82 | **< 0.001*** ** | **0.002** ** |
| 3 (12 months from Baseline) | con.SuperiorFrontal | con.CaudalMiddleFrontal | 0.92 | **< 0.001*** ** | **< 0.001*** ** |
| | con.SuperiorParietal | con.Precuneus | 0.90 | **< 0.001*** ** | **0.001*** ** |
| | con.SuperiorFrontal | con.RostralMiddleFrontal | 0.90 | **< 0.001*** ** | **0.001*** ** |
| | con.TransverseTemporal | con.SuperiorParietal | 0.89 | **< 0.001*** ** | **0.002** ** |
| | con.Precuneus | con.ParsOrbitalis | 0.88 | **< 0.001*** ** | **0.002** ** |
| | con.SuperiorParietal | con.ParsOrbitalis | 0.87 | **< 0.001*** ** | **0.003** ** |
| | con.Supramarginal | con.ParsOpercularis | 0.87 | **< 0.001*** ** | **0.004** ** |
| | ip.LateralOrbitofrontal | con.CaudalMiddleFrontal | 0.86 | **< 0.001*** ** | **0.004** ** |
| | ip.LateralOrbitofrontal | con.RostralMiddleFrontal | 0.85 | **< 0.001*** ** | **0.007** ** |
| | ip.Insula | con.RostralAnteriorCingulate | 0.84 | **< 0.001*** ** | **0.008** ** |
| | con.ParsOpercularis | con.InferiorParietal | 0.84 | **< 0.001*** ** | **0.009** ** |
| | con.Supramarginal | con.InferiorParietal | 0.83 | **< 0.001*** ** | **0.009** ** |

*Note.* The top 12 significant region pairs (i.e., edges) of Sessions 2 and 3 are displayed. The adjusted p-values displayed are following FDR correction using the BH procedure. All *p*-values were rounded to three decimal places. * $p < 0.05$, ** $p < 0.01$, *** $p < 0.001$. CON, contralesional; IP, ipsilesional.
